# Added value of serum proteins to clinical and ultrasound information in predicting the risk of malignancy in ovarian tumors

**DOI:** 10.1101/2024.03.14.24304282

**Authors:** A Coosemans, J Ceusters, C Landolfo, T Baert, W Froyman, R Heremans, G Thirion, S Claes, J Oosterlynck, R Wouters, A Vankerckhoven, F Moro, F Mascilini, A Neumann, AS Van Rompuy, D Schols, J Billen, T Van Gorp, I Vergote, T Bourne, C Van Holsbeke, V Chiappa, G Scambia, A Testa, D Fischerova, D Timmerman, B Van Calster

## Abstract

**Background:** The ADNEX model (Assessment of Different NEoplasias in the adnexa) is the best performing model to predict the risk of malignancy (binary) and type of malignancy (multiclass) in ovarian tumors. The immune system plays a role in the onset and progression of ovarian cancer. Preliminary research has suggested that immune-related biomarkers can help in the discrimination of ovarian tumors. We aimed to assess which proteins have the most additional diagnostic value in addition to ADNEX’ clinical and ultrasound predictors.

**Materials and methods:** In this exploratory diagnostic study, 1086 patients with an adnexal mass scheduled for surgery were consecutively enrolled at five oncology centers and one non-oncology center in Belgium, Italy, Czech Republic and United Kingdom between 2015 and 2019. The quantification of 33 serum proteins was carried out preoperatively, using multiplex high throughput immunoassays (Luminex) and electrochemiluminescence immuno-assay (ECLIA). Logistic regression analysis was performed for ADNEX’ clinical and ultrasound predictors alone (age, maximum diameter of lesion, proportion of solid tissue, presence of >10 cyst locules, number of papillary projections, acoustic shadows and ascites) and after adding proteins. We reported the AUC for benign vs malignant, Polytomous Discrimination Index (PDI; a multiclass AUC) and pairwise AUCs for pairs of tumor types. AUCs were corrected for optimism using bootstrapping.

**Results:** After applying exclusion criteria, 932/1086 patients were eligible for analysis (474 benign, 135 borderline, 84 stage I primary invasive cancer, 208 stage II-IV primary invasive cancer, 31 secondary metastatic invasive tumors). ADNEX predictors alone had an AUC of 0.909 (95% CI 0.894-0.929) to discriminate benign from malignant tumors, and a PDI of 0.532 (0.510-0.589). HE4 yielded the highest increase in AUC (+0.026), followed by CA125 (+0.017). CA125 yielded the highest increase in PDI (+0.049), followed by HE4 (+0.036). Whereas CA125 mainly improved pairwise AUCs between different types of invasive tumors (increases between 0.020-0.165 over ADNEX alone), HE4 mainly improved pairwise AUCs for benign tumors versus stage I (+0.022) and benign tumors versus stage II-IV ovarian cancers (+0.028). CA72.4 might be useful to distinguishing secondary metastatic tumors from benign, borderline, and stage I tumors. CA15.3 might be useful to discriminate borderline tumors from stage I and stage II-IV tumors. Distinguishing stage I and borderline tumors (AUCs ≤ 0.72) and stage I and secondary metastatic tumors (AUCs ≤ 0.76) remained difficult after adding proteins.

**Conclusions:** CA125 had the highest added value over clinical and ultrasound predictors to distinguish between the five tumor types, followed by HE4. In addition, CA72.4 and CA15.3 may further improve discrimination but findings for these proteins should be confirmed. The immune-related proteins were in general not able to discriminate the groups.

## Introduction

Approximately 1.1 percent of women will be diagnosed with ovarian cancer at some point during their lifetime^1^. Diagnostic accuracy is of utmost importance because patient management will depend on it. A benign ovarian cyst does not need intervention in the majority of cases, a stage I ovarian cancer needs to be handled differently compared to advanced stage disease or a borderline tumor, and treatment of an ovarian metastasis differs from primary ovarian cancer^2–4^. The IOTA group has optimized the diagnostic potential of transvaginal ultrasound by using the ADNEX risk prediction model.^5^ ADNEX estimates the risk of five tumor types: benign, borderline, stage I primary invasive, stage II-IV primary invasive, and secondary invasive (metastasis to the ovary) tumors. ADNEX has an AUC of 0.93 to discriminate between benign and any type of malignant tumors in patients scheduled for surgery or expectant management.^6–7^ The protein biomarker CA125 is an optional predictor in ADNEX. Using the version with CA125 mainly improves the discrimination of stage II-IV invasive tumors from stage I invasive and secondary metastatic tumors.^5,7^ Nevertheless, the AUC between some malignant subtypes is around 0.7-0.8: a meta-analysis reported AUCs of 0.72 for borderline versus stage I primary invasive tumors, and 0.78-0.82 for discrimination between invasive subtypes.^7^ Next to CA125, HE4 (Human Epididymis Protein 4) is also often used. Based on a recent systematic review, however, HE4 does not outperform CA125 to discriminate benign from malignant tumors.^8^

Over the past two decades, it has become clear that the immune system is an important factor in the onset and progression of cancer.^9^ The focus for ovarian cancer immune biology has been on the adaptive immune system. CD8^+^ T cells at diagnosis are positively associated with prognosis, while regulatory T cells are negatively associated.^10,11^ However, more and more reports have now indicated that the immune system is much more complex in ovarian cancer and that the importance of the innate immune system cannot be underestimated.^12,13^ Our previous work suggested that the immune system, both at the cellular and the protein level, may be used as a biomarker to discriminate benign from malignant ovarian tumors.^14,15^

The goal of this study was to evaluate if a combination of ultrasound and immune-related parameters could result in a further improvement of performance of the ADNEX model.

## Materials and Methods

### Study design and participants

In this exploratory prospective cohort study, we recruited patients who presented with an adnexal (i.e. ovarian/paraovarian, tubal/paratubal) tumor. They were consecutively enrolled at six hospitals between June 2015 and December 2019: the University Hospitals Leuven (Leuven, Belgium), Università Cattolica del Sacro Cuore (Rome, Italy), General Faculty Hospital of the Charles University (Prague, Czech Republic), Queen Charlotte’s and Chelsea Hospital (London, UK), Ziekenhuis Oost-Limburg (Genk, Belgium), and Istituto Nazionale dei Tumore (Milan, Italy). The Research Ethics Committee of the University Hospitals Leuven acted as central ethics committee and approved the study (S51375, S59207). Ethics approval was also obtained at all other local ethics committees. Written informed consent was required before recruitment. All patients underwent a preoperative standardized transvaginal ultrasound scan according to the International Ovarian Tumor Analysis (IOTA) methodology^16^ in the framework of IOTA5 and IOTA7 studies. All patients were selected for surgery by the managing clinician. Exclusion criteria were the following: (1) age less than 18 years old, (2) pregnancy, (3) refusal of preoperative transvaginal ultrasonography and/or blood sample, (4) known simultaneous and/or previous malignancies within five years, (5) simultaneous autoimmune disease and/or treatment with immune modulating drugs, (6) infectious serology (i.e. HIV, Hepatitis B, Hepatitis C), (7) denial or withdrawal of the written informed consent, and (8) surgery more than 120 days after the ultrasound examination. Data cleaning was performed by a team of biostatisticians and ultrasound examiners. Data cleaning included sending queries to participating hospitals to retrieve missing information or to correct inconsistencies. We report the study according to the REMARK (Reporting Recommendations for Tumor Marker Prognostic Studies) guideline.^17^ Of the patient samples collected in Leuven, 254 of them were used in a biomarker paper, published earlier by our group in 2020.^15^

### Serum samples

Serum from a blood sample (BD Vacutainer SST II advance), taken before surgery from each patient, was isolated following a specific protocol (**in Supplementary Appendix 1**). Serum was subsequently stored in aliquots at -80°C. Samples were shipped to the Laboratory of Tumor Immunology and Immunotherapy at KU Leuven, where the assays were performed. After thawing, aliquots were used for analysis (i.e. defrosted samples were not refrozen for additional analyses).

### Protein analysis

Circulating levels of 33 proteins were evaluated **(Table 1)**. Proteins were measured with Luminex® assay and electrochemiluminescence immuno-assay (ECLIA), according to the manufacturers’ instructions. For Luminex™ technology, assays were performed using aliquots of 150 µL of serum. Customized Procartaplex™ Immunoassays Kits were purchased (Life Technologies, Merelbeke, Belgium) to determine 23 proteins. The procedure used has been described earlier by our group.^15^ For ECLIA (Roche Diagnostics), 500 µl of serum was required. Kits were obtained from Roche Diagnostics to determine ten proteins. Both assay procedures have been described earlier by our group.^15^

**Table 1.** Overview on proteins measured in serum.

| Protein | Assay | At/below lower limit of detection, n (%) | Missing values, n (%) |
| --- | --- | --- | --- |
| Arginase | Luminex | 248 (27%) | 86 (9%) |
| CCL11 | Luminex | 1 (0.1%) | 86 (9%) |
| CCL24 | Luminex | 0 (0%) | 86 (9%) |
| Galectin-3 | Luminex | 377 (40%) | 86 (9%) |
| IL-6 | ECLIA | 98 (11%) | 41 (4%) |
| IL-10 | Luminex | 56 (6%) | 86 (9%) |
| IL-12p70 | Luminex | 722 (77%) | 73 (8%) |
| IL-17F | Luminex | 785 (84%) | 87 (9%) |
| IP-10/CXCL10 | Luminex | 11 (1%) | 73 (8%) |
| TGF-β/LAP | Luminex | 316 (34%) | 73 (8%) |
| MCP-1/CCL2 | Luminex | 411 (44%) | 73 (8%) |
| MDC/CCL22 | Luminex | 8 (0.9%) | 73 (8%) |
| MIG/CXCL9 | Luminex | 126 (14%) | 86 (9%) |
| MMP-1 | Luminex | 1 (0.1%) | 338 (36%) |
| MMP-7 | Luminex | 2 (0.2%) | 86 (9%) |
| MMP-8 | Luminex | 16 (2%) | 338 (36%) |
| MMP-9 | Luminex | 2 (0.2%) | 86 (9%) |
| MMP-12 | Luminex | 225 (24%) | 338 (36%) |
| MMP-13 | Luminex | 315 (34%) | 338 (36%) |
| OPN | Luminex | 216 (23%) | 86 (9%) |
| PIGF | ECLIA | 4 (0.4%) | 45 (5%) |
| proGRP | ECLIA | 0 (0%) | 55 (6%) |
| CCL5 | Luminex | 10 (1%) | 73 (8%) |
| SAA | Luminex | 15 (2%) | 86 (9%) |
| SDF-1α/CXCL12a | Luminex | 16 (2%) | 73 (8%) |
| sFLT1 | ECLIA | 0 (0%) | 40 (4%) |
| VEGF-A | Luminex | 7 (0.8%) | 73 (8%) |
| CA125 | ECLIA | 1 (0.1%) | 38 (4%) |
| HE4 | ECLIA | 0 (0%) | 38 (4%) |
| CA72.4 | ECLIA | 301 (32%) | 38 (4%) |
| CA19.9 | ECLIA | 78 (8%) | 38 (4%) |
| CA15.3 | ECLIA | 0 (0%) | 37 (4%) |
| CEA | ECLIA | 3 (0.3%) | 37 (4%) |

### Reference standard

The reference standard was the histopathological diagnosis by the local pathologist after surgical removal of the adnexal mass. Tumors were classified according to the WHO (World Health Organization) classification of tumors and malignant lesions were staged according to the FIGO (International Federation of Gynecology and Obstetrics) criteria.^18^

### ADNEX model

The ADNEX model is a multinomial logistic regression model based on three clinical predictors (age, serum CA125, type of center) and six ultrasound predictors (maximal diameter of the lesion, proportion of solid tissue, more than ten cyst locules, number of papillary projections, acoustic shadowing, and presence of ascites).^5^ Type of center refers to oncology referral units vs other units. In this study, only one center (Genk, Belgium) was not an oncology center. ADNEX estimates the risk of five tumor types: benign, borderline, stage I and stage II-IV primary invasive ovarian cancer, and secondary metastasis. One minus the probability of a benign tumor equals the estimated risk of malignancy.

### Statistical analysis

The 33 proteins considered as parameters were log-transformed (log with base 2) for all statistical analyses. Missing values (due to technical issues and/or insufficient serum sample) and values out of range were addressed using single stochastic imputation based on ‘multivariate imputation by chained equations’ (mice).^19^ The protein levels were imputed using predictive mean matching regression. Imputation was based on the predictors in the ADNEX model (except type of center), all protein measurements, and outcome.

The unadjusted discriminative ability of each protein was calculated using univariable areas under the receiver operating characteristic curves (AUCs), with 95% confidence interval (CI) based on the logit transform method.^20^ Comparisons were performed between benign masses and all malignant tumors, as well as between the five tumor types: benign, borderline, stage I invasive, stage II-IV invasive, and secondary metastasis.

To investigate the added diagnostic value of the proteins for predicting the risk of malignancy, we first fitted a binary logistic regression model of the outcome (benign vs any type of malignancy) on seven clinical and ultrasound predictors from the ADNEX model: age, maximum diameter of lesion, proportion of solid tissues, presence of more than 10 cyst locules, number of papillary projections, presence of acoustic shadows, and presence of ascites. Type of center was not used because all but one centers were oncology referral centers and contributed 92% of patients. Then, we fitted similar models with the seven ADNEX predictors plus (1) each protein separately, (2) CA125 and HE4, and (3) a variable selection of proteins only using the lasso (least absolute shrinkage and selection operator). The lasso selection was based on a model with the seven ADNEX predictors and all proteins in which a lasso-penalty was applied to the proteins but not to the ADNEX predictors.^21,22^ For all these models, the AUC was calculated. To investigate the added value for predicting the risk of malignant subtypes, similar analyses were performed using multinomial logistic regression for the five tumor types predicted by ADNEX. For the multinomial models, the Polytomous Discrimination Index (PDI) was calculated as a multiclass AUC extension, as well as pairwise AUCs using the conditional risk method.^23,24^ Internal validation was performed using bootstrapping with 200 resamples to correct AUCs and PDIs for optimism. All regression models included Firth’s correction.^25^

The sensitivity and specificity for CA125 and HE4 at common cut-offs (35 U/mL for CA125 and 70 pmol/L for HE4^8^) were calculated for each center. Overall values were based on random-effects bivariate logit-normal model. Also, an overall estimate while ignoring the multicenter nature of the data was calculated, using 95% confidence intervals [CI] based on Wilson’s score method.^26^

Two sensitivity analyses were performed for the univariable AUCs: (1) by using meta-analysis of center-specific AUCs, and (2) after using multiple imputation (100 imputations) for the missing values.

All statistical analyses were performed in R version 4.1.0, using packages mice, auRoc, logistf, glmnet, mada and metafor.

## Results

### Patient characteristics

In total, 1090 patients were enrolled in the study. After applying the exclusion criteria, the final cohort consisted of 932 patients with an ovarian mass, collected at six centers (**Figure 1**). Among these patients, 474 (51%) had a benign and 458 (49%) a malignant adnexal tumor. Among the malignant tumors, there were 135 (14%) borderline tumors, 84 (9%) stage I primary invasive tumors, 208 (22%) stage II-IV primary invasive tumors and 31 (3%) secondary metastatic tumors. The patients had a median age of 54 years (IQR (intrquartile range) 42-64, range 18-88) and 58% of the patients were postmenopausal (**Table 2**).

**Figure 1.**
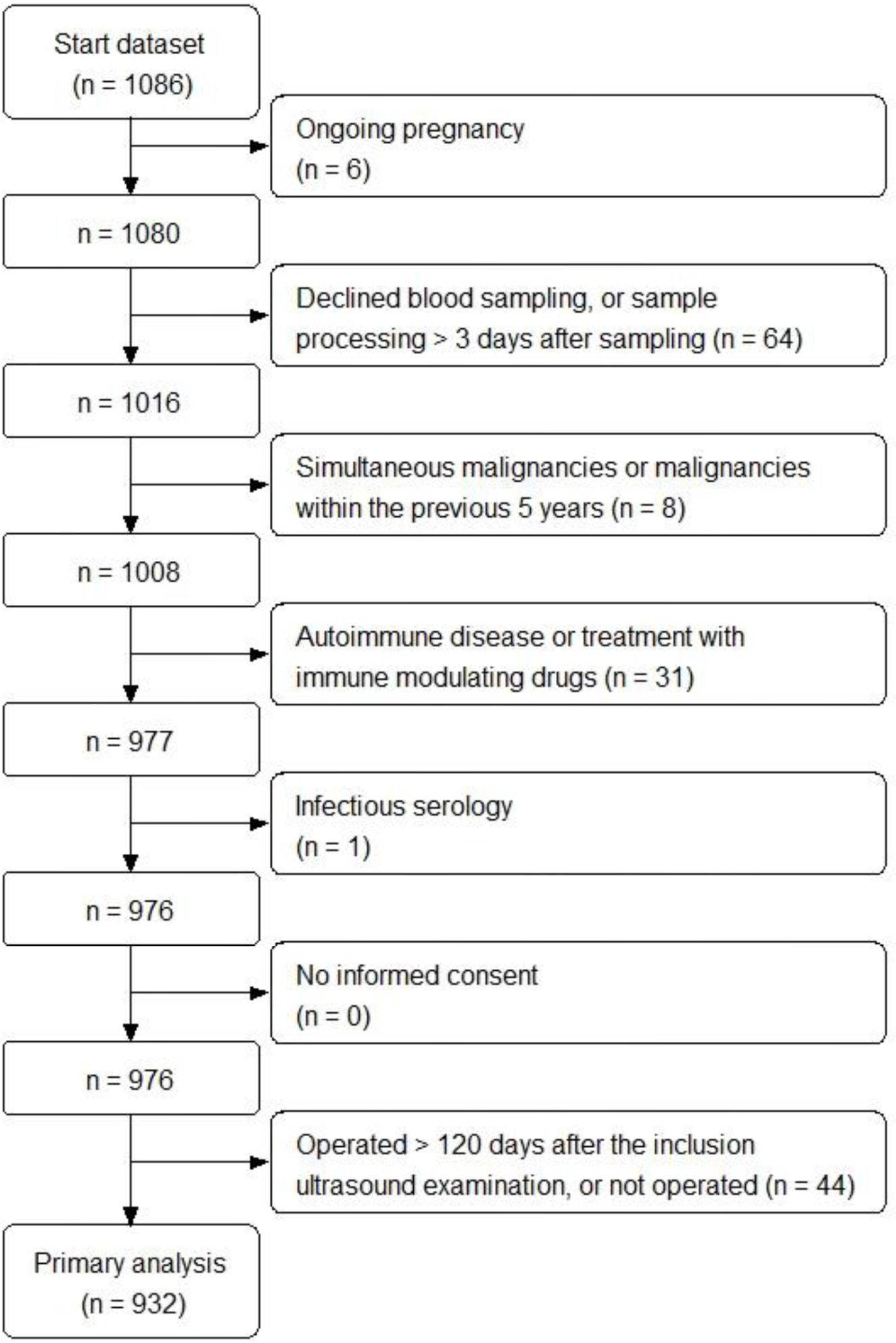
Flowchart of the patients included in the analysis.

**Table 2.** Descriptive statistics (n = 932).

| Variable | Median (IQR) (range), or n (%) |
| --- | --- |
| Patient age at recruitment (years) | 54 (42-64) (18-88) |
| Postmenopausal | 536 (58%) |
| Presence of solid components | 636 (68%) |
| Maximum diameter of lesion (mm) | 75 (49-117) (7-459) |
| Largest diameter of largest solid component* | 45 (24-71) (3-400) |
| Number of papillary projections |  |
| 0 | 655 (70%) |
| 1 | 122 (13%) |
| 2 | 37 (4%) |
| 3 | 23 (2%) |
| 4 | 95 (10%) |
| Bilateral Masses | 241 (26%) |
| More than 10 cyst locules | 143 (15%) |
| Acoustic shadows | 338 (36%) |
| Ascites | 141 (15%) |
| Ultrasound examiner's subjective impression |  |
| Probably benign | 207 (22%) |
| Certainly benign | 138 (15%) |
| Probably malignant | 160 (17%) |
| Certainly malignant | 309 (33%) |
| Uncertain | 118 (13%) |
| Outcome |  |
| Benign | 474 (51%) |
| Borderline | 135 (14%) |
| Serous | 72 (8%) |
| Non-serous | 63 (7%) |
| FIGO Stage I invasive | 84 (9%) |
| High grade serous epithelial | 13 (1%) |
| Low grade serous epithelial | 2 (<1%) |
| Non-serous epithelial | 54 (6%) |
| Non-epithelial | 15 (26%) |
| FIGO Stage II – IV invasive | 208 (22%) |
| High grade serous epithelial | 162 (17%) |
| Low grade serous epithelial | 14 (2%) |
| Non-serous epithelial | 28 (3%) |
| Non-epithelial | 4 (<1%) |
| Metastatic | 31 (3%) |

\* Only for tumors with a solid component
Percentages may not sum to 100 due to rounding.
IQR, interquartile range

### Protein characteristics

For the analysis, we excluded eight of the 33 investigated proteins. Four proteins were excluded because a large majority of samples were at or below the lower limit of detection (IL-12p70, IL17F, MCP-1, Galectin-3) (**Supplementary** Figure 1). The other four proteins were omitted because these proteins were not measured from the start (Procartaplex composition was changed by Life Technologies during the analysis of the study), resulting in a large number of missing values (MMP1, MMP8, MMP12 and MMP13). **Figure 2** shows the distribution of the protein data according to pathological analysis, after logarithmic transformation. The correlation with the variables in the ADNEX model is shown in **Supplementary Table 1**.

**Figure 2.**
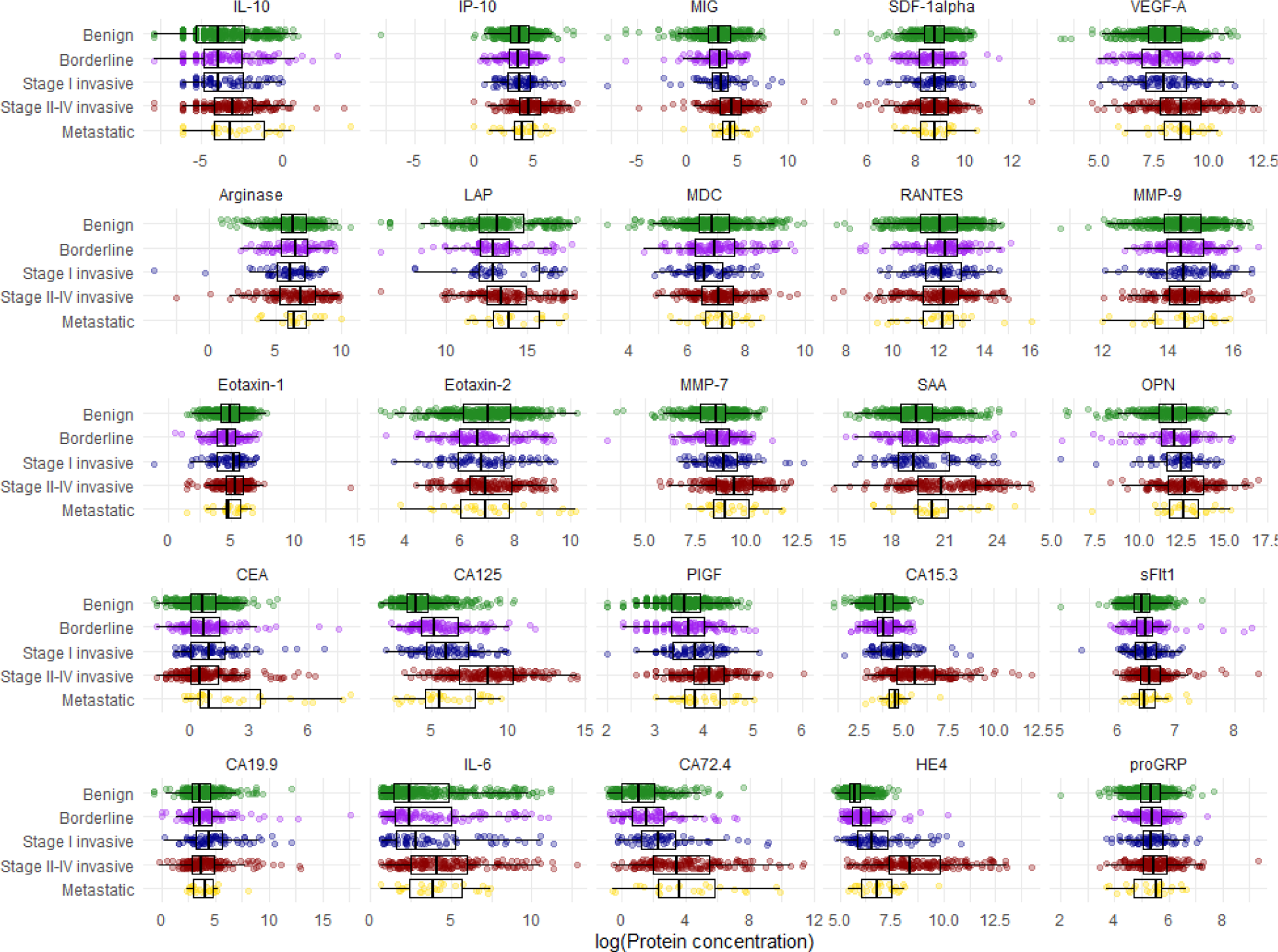
The distribution of each investigated protein according to histology. Data after logarithmic transformation (log2 transformation) are presented.

### Univariable AUCs

The highest univariable AUCs were observed for CA125 (AUC of 0.84), HE4 (0.82), CA72.4 (0.75) and CA15.3 (0.71) (**Table 3**, **Supplementary** Figure 2) to discriminate benign from malignant tumors. If the variability in performance between the six hospitals was taken into account (first sensitivity analysis), CA125, HE4, CA72.4 and CA15.3 still had the highest AUC values (**Supplementary Figure & Table 3**). Interestingly, the performance of some proteins (MDC, MMP-7, PlGF, sFlt1 and CA72.4) showed substantial heterogeneity between the different hospitals. When the missing values were multiply imputed (second sensitivity analysis), the same overall results were obtained (**Supplementary Figure & Table 4**).

**Table 3.** Univariable AUCs (95%Confidence Intervals).

| Protein | Benign vs Malignant | Benign vs Borderline | Benign vs Stage I | Benign vs Stage II-IV | Benign vs Metastatic | Borderline vs Stage I | Borderline vs Stage II-IV | Borderline vs Metastatic | Stage I vs Stage II-IV | Stage I vs Metastatic | Stage II-IV vs Metastatic |
| --- | --- | --- | --- | --- | --- | --- | --- | --- | --- | --- | --- |
| CA125 | 0.84 (0.81; 0.86) | 0.74 (0.68; 0.78) | 0.78 (0.72; 0.83) | 0.95 (0.93; 0.96) | 0.72 (0.62; 0.81) | 0.58 (0.51; 0.66) | 0.85 (0.81; 0.89) | 0.52 (0.41; 0.63) | 0.80 (0.74; 0.85) | 0.56 (0.44; 0.67) | 0.83 (0.74; 0.89) |
| HE4 | 0.82 (0.79; 0.85) | 0.64 (0.59; 0.69) | 0.78 (0.72; 0.83) | 0.96 (0.94; 0.97) | 0.83 (0.74; 0.89) | 0.67 (0.59; 0.74) | 0.92 (0.88; 0.95) | 0.73 (0.62; 0.81) | 0.81 (0.75; 0.86) | 0.54 (0.42; 0.65) | 0.82 (0.73; 0.89) |
| CA72.4 | 0.75 (0.68; 0.81) | 0.60 (0.55; 0.66) | 0.72 (0.66; 0.77) | 0.85 (0.81; 0.88) | 0.83 (0.75; 0.90) | 0.62 (0.55; 0.70) | 0.77 (0.72; 0.82) | 0.77 (0.66; 0.85) | 0.67 (0.60; 0.73) | 0.68 (0.56; 0.78) | 0.52 (0.41; 0.63) |
| CA15.3 | 0.71 (0.67; 0.74) | 0.51 (0.46; 0.57) | 0.66 (0.60; 0.72) | 0.88 (0.85; 0.91) | 0.70 (0.60; 0.79) | 0.67 (0.59; 0.74) | 0.88 (0.84; 0.91) | 0.71 (0.60; 0.80) | 0.77 (0.70; 0.82) | 0.51 (0.39; 0.63) | 0.78 (0.68; 0.85) |
| PIGF | 0.65 (0.59; 0.71) | 0.53 (0.47; 0.58) | 0.60 (0.54; 0.67) | 0.76 (0.72; 0.80) | 0.70 (0.60; 0.78) | 0.57 (0.49; 0.64) | 0.73 (0.67; 0.78) | 0.66 (0.55; 0.76) | 0.67 (0.60; 0.73) | 0.60 (0.48; 0.71) | 0.57 (0.46; 0.67) |
| MMP-7 | 0.64 (0.58; 0.69) | 0.56 (0.51; 0.62) | 0.61 (0.55; 0.68) | 0.74 (0.70; 0.78) | 0.68 (0.57; 0.76) | 0.56 (0.48; 0.63) | 0.69 (0.63; 0.74) | 0.63 (0.52; 0.73) | 0.63 (0.56; 0.70) | 0.57 (0.45; 0.69) | 0.55 (0.45; 0.66) |
| IL-6 | 0.64 (0.60; 0.68) | 0.54 (0.49; 0.59) | 0.56 (0.49; 0.62) | 0.69 (0.65; 0.73) | 0.67 (0.57; 0.76) | 0.52 (0.44; 0.60) | 0.66 (0.60; 0.71) | 0.64 (0.53; 0.74) | 0.63 (0.56; 0.70) | 0.60 (0.48; 0.71) | 0.54 (0.43; 0.64) |
| IP-10 | 0.62 (0.58; 0.65) | 0.51 (0.45; 0.56) | 0.52 (0.46; 0.59) | 0.71 (0.67; 0.75) | 0.58 (0.47; 0.68) | 0.53 (0.45; 0.61) | 0.72 (0.66; 0.77) | 0.59 (0.47; 0.69) | 0.68 (0.61; 0.75) | 0.56 (0.44; 0.67) | 0.63 (0.52; 0.72) |
| MIG | 0.61 (0.58; 0.65) | 0.52 (0.46; 0.57) | 0.51 (0.45; 0.58) | 0.69 (0.65; 0.74) | 0.70 (0.60; 0.79) | 0.50 (0.42; 0.58) | 0.70 (0.64; 0.75) | 0.71 (0.61; 0.80) | 0.68 (0.61; 0.75) | 0.70 (0.58; 0.79) | 0.51 (0.40; 0.62) |
| SAA | 0.61 (0.57; 0.65) | 0.53 (0.47; 0.58) | 0.51 (0.45; 0.58) | 0.72 (0.68; 0.76) | 0.67 (0.57; 0.76) | 0.51 (0.43; 0.59) | 0.69 (0.64; 0.75) | 0.64 (0.53; 0.74) | 0.69 (0.62; 0.75) | 0.64 (0.52; 0.74) | 0.56 (0.45; 0.66) |
| OPN | 0.61 (0.57; 0.65) | 0.52 (0.46; 0.57) | 0.58 (0.51; 0.64) | 0.65 (0.60; 0.69) | 0.70 (0.60; 0.78) | 0.59 (0.51; 0.66) | 0.66 (0.60; 0.71) | 0.70 (0.59; 0.79) | 0.57 (0.50; 0.64) | 0.61 (0.49; 0.72) | 0.54 (0.43; 0.64) |
| IL-10 | 0.60 (0.55; 0.65) | 0.52 (0.47; 0.58) | 0.51 (0.45; 0.58) | 0.63 (0.58; 0.67) | 0.62 (0.52; 0.72) | 0.51 (0.43; 0.59) | 0.61 (0.54; 0.67) | 0.61 (0.49; 0.71) | 0.62 (0.55; 0.69) | 0.63 (0.51; 0.73) | 0.51 (0.40; 0.61) |
| sFlt1 | 0.60 (0.54; 0.65) | 0.57 (0.51; 0.62) | 0.57 (0.50; 0.63) | 0.66 (0.61; 0.70) | 0.62 (0.51; 0.71) | 0.51 (0.43; 0.59) | 0.59 (0.53; 0.65) | 0.55 (0.44; 0.66) | 0.58 (0.51; 0.65) | 0.54 (0.42; 0.66) | 0.54 (0.43; 0.65) |
| Arginase | 0.59 (0.55; 0.62) | 0.54 (0.49; 0.60) | 0.56 (0.49; 0.62) | 0.61 (0.56; 0.65) | 0.55 (0.44; 0.65) | 0.51 (0.43; 0.58) | 0.56 (0.50; 0.62) | 0.50 (0.39; 0.61) | 0.56 (0.49; 0.63) | 0.51 (0.39; 0.63) | 0.56 (0.45; 0.66) |
| VEGF-A | 0.58 (0.54; 0.62) | 0.52 (0.47; 0.58) | 0.51 (0.45; 0.58) | 0.66 (0.62; 0.71) | 0.66 (0.56; 0.75) | 0.53 (0.45; 0.61) | 0.68 (0.62; 0.73) | 0.67 (0.56; 0.77) | 0.65 (0.58; 0.71) | 0.64 (0.52; 0.74) | 0.52 (0.41; 0.63) |
| LAP | 0.57 (0.53; 0.61) | 0.51 (0.45; 0.56) | 0.50 (0.44; 0.57) | 0.63 (0.59; 0.68) | 0.65 (0.55; 0.74) | 0.51 (0.43; 0.58) | 0.63 (0.57; 0.69) | 0.65 (0.54; 0.75) | 0.63 (0.56; 0.70) | 0.64 (0.52; 0.75) | 0.51 (0.40; 0.62) |
| proGRP | 0.57 (0.54; 0.61) | 0.54 (0.48; 0.59) | 0.54 (0.47; 0.61) | 0.59 (0.55; 0.64) | 0.54 (0.44; 0.64) | 0.51 (0.43; 0.58) | 0.56 (0.50; 0.62) | 0.50 (0.39; 0.61) | 0.55 (0.48; 0.62) | 0.50 (0.38; 0.62) | 0.55 (0.44; 0.65) |
| MDC | 0.56 (0.51; 0.61) | 0.52 (0.47; 0.58) | 0.58 (0.52; 0.65) | 0.55 (0.51; 0.60) | 0.58 (0.47; 0.68) | 0.60 (0.52; 0.68) | 0.53 (0.46; 0.59) | 0.55 (0.43; 0.66) | 0.64 (0.56; 0.70) | 0.66 (0.54; 0.76) | 0.52 (0.42; 0.63) |
| Eotaxin-1 | 0.56 (0.53; 0.60) | 0.55 (0.50; 0.61) | 0.54 (0.48; 0.61) | 0.61 (0.56; 0.65) | 0.57 (0.46; 0.67) | 0.60 (0.52; 0.67) | 0.66 (0.60; 0.72) | 0.62 (0.51; 0.72) | 0.56 (0.49; 0.64) | 0.52 (0.40; 0.63) | 0.55 (0.44; 0.66) |
| CEA | 0.56 (0.52; 0.60) | 0.53 (0.48; 0.59) | 0.56 (0.50; 0.63) | 0.51 (0.47; 0.56) | 0.67 (0.56; 0.76) | 0.53 (0.45; 0.60) | 0.54 (0.48; 0.60) | 0.64 (0.53; 0.74) | 0.57 (0.49; 0.64) | 0.61 (0.49; 0.72) | 0.67 (0.56; 0.76) |
| CA19.9 | 0.56 (0.53; 0.60) | 0.53 (0.48; 0.59) | 0.63 (0.56; 0.69) | 0.53 (0.48; 0.58) | 0.53 (0.42; 0.63) | 0.59 (0.51; 0.67) | 0.50 (0.44; 0.57) | 0.51 (0.40; 0.62) | 0.59 (0.52; 0.66) | 0.61 (0.49; 0.72) | 0.51 (0.40; 0.61) |
| MMP-9 | 0.55 (0.51; 0.58) | 0.51 (0.45; 0.56) | 0.54 (0.47; 0.61) | 0.53 (0.49; 0.58) | 0.55 (0.44; 0.65) | 0.53 (0.46; 0.61) | 0.53 (0.47; 0.59) | 0.56 (0.45; 0.67) | 0.51 (0.43; 0.58) | 0.58 (0.46; 0.69) | 0.57 (0.46; 0.68) |
| SDF-1α | 0.53 (0.49; 0.57) | 0.51 (0.45; 0.56) | 0.50 (0.44; 0.57) | 0.52 (0.47; 0.56) | 0.52 (0.41; 0.62) | 0.50 (0.43; 0.58) | 0.52 (0.46; 0.58) | 0.52 (0.41; 0.63) | 0.52 (0.44; 0.59) | 0.51 (0.40; 0.63) | 0.50 (0.39; 0.61) |
| Eotaxin-2 | 0.53 (0.49; 0.57) | 0.54 (0.48; 0.59) | 0.53 (0.46; 0.59) | 0.52 (0.48; 0.57) | 0.53 (0.42; 0.63) | 0.51 (0.43; 0.59) | 0.57 (0.50; 0.63) | 0.56 (0.45; 0.67) | 0.55 (0.48; 0.63) | 0.56 (0.44; 0.67) | 0.50 (0.39; 0.61) |
| RANTES | 0.52 (0.48; 0.56) | 0.52 (0.47; 0.58) | 0.51 (0.44; 0.57) | 0.53 (0.48; 0.58) | 0.59 (0.48; 0.68) | 0.52 (0.44; 0.59) | 0.51 (0.45; 0.57) | 0.61 (0.50; 0.72) | 0.53 (0.45; 0.60) | 0.59 (0.47; 0.70) | 0.62 (0.51; 0.71) |

**Table 4.** Univariable AUCs (95% Confidence interval) for the comparison of different types of borderlines (BOT) and stage I invasive tumors.

| Protein | All BOT vs serous<br>epithelial Stage I | Serous BOT vs<br>serous epithelial Stage<br>I | Non-serous BOT vs<br>non-serous Stage I | All BOT vs<br>non-epithelial Stage I |
| --- | --- | --- | --- | --- |
| CA125 | 0.66 (0.50-0.80) | 0.62 (0.45-0.77) | 0.62 (0.52-0.72) | 0.50 (0.35-0.65) |
| HE4 | 0.81 (0.65-0.90) | 0.82 (0.66-0.91) | 0.66 (0.55-0.75) | 0.51 (0.36-0.66) |
| CA72.4 | 0.73 (0.57-0.85) | 0.77 (0.60-0.88) | 0.61 (0.50-0.70) | 0.59 (0.44-0.73) |
| CA15.3 | 0.85 (0.71-0.93) | 0.89 (0.75-0.96) | 0.63 (0.52-0.72) | 0.56 (0.40-0.70) |
| PIGF | 0.53 (0.37-0.68) | 0.54 (0.37-0.70) | 0.56 (0.45-0.66) | 0.52 (0.37-0.67) |
| MMP-7 | 0.55 (0.39-0.70) | 0.58 (0.41-0.74) | 0.57 (0.46-0.67) | 0.63 (0.47-0.76) |
| IL-6 | 0.51 (0.35-0.67) | 0.50 (0.34-0.67) | 0.55 (0.44-0.65) | 0.53 (0.38-0.68) |
| IP-10 | 0.69 (0.52-0.81) | 0.68 (0.51-0.82) | 0.52 (0.42-0.62) | 0.56 (0.40-0.70) |
| MIG | 0.66 (0.49-0.79) | 0.65 (0.48-0.79) | 0.52 (0.42-0.63) | 0.52 (0.37-0.66) |
| SAA | 0.52 (0.36-0.68) | 0.52 (0.35-0.68) | 0.50 (0.40-0.61) | 0.57 (0.41-0.71) |
| OPN | 0.57 (0.41-0.72) | 0.59 (0.42-0.74) | 0.57 (0.46-0.66) | 0.62 (0.47-0.76) |
| IL-10 | 0.53 (0.37-0.69) | 0.55 (0.38-0.71) | 0.55 (0.44-0.65) | 0.50 (0.35-0.65) |
| sFlt1 | 0.51 (0.35-0.67) | 0.50 (0.34-0.67) | 0.50 (0.40-0.61) | 0.51 (0.36-0.66) |
| Arginase | 0.62 (0.45-0.76) | 0.63 (0.45-0.77) | 0.53 (0.43-0.63) | 0.53 (0.38-0.68) |
| VEGF-A | 0.56 (0.40-0.71) | 0.57 (0.39-0.72) | 0.53 (0.42-0.63) | 0.53 (0.38-0.67) |
| LAP | 0.66 (0.49-0.79) | 0.63 (0.46-0.78) | 0.50 (0.40-0.61) | 0.52 (0.37-0.67) |
| proGRP | 0.54 (0.38-0.69) | 0.51 (0.34-0.67) | 0.50 (0.40-0.60) | 0.56 (0.41-0.70) |
| MDC | 0.63 (0.46-0.77) | 0.66 (0.49-0.80) | 0.56 (0.46-0.66) | 0.64 (0.48-0.77) |
| Eotaxin-1 | 0.73 (0.57-0.85) | 0.73 (0.56-0.85) | 0.61 (0.51-0.71) | 0.55 (0.40-0.70) |
| CEA | 0.59 (0.43-0.74) | 0.52 (0.36-0.69) | 0.51 (0.40-0.61) | 0.58 (0.43-0.72) |
| CA19.9 | 0.57 (0.41-0.72) | 0.51 (0.34-0.67) | 0.57 (0.46-0.67) | 0.52 (0.36-0.66) |
| MMP-9 | 0.55 (0.39-0.70) | 0.58 (0.41-0.74) | 0.60 (0.50-0.70) | 0.50 (0.35-0.65) |
| SDF-1α | 0.61 (0.45-0.76) | 0.59 (0.42-0.74) | 0.51 (0.41-0.62) | 0.53 (0.37-0.67) |
| Eotaxin-2 | 0.67 (0.50-0.80) | 0.66 (0.49-0.80) | 0.53 (0.42-0.63) | 0.66 (0.50-0.79) |
| RANTES | 0.64 (0.47-0.78) | 0.65 (0.47-0.79) | 0.51 (0.41-0.62) | 0.53 (0.38-0.68) |

**Supplementary Table 4** shows the results for sensitivity and specificity of CA125 and HE4 at commonly used cut-offs (35 U/mL and 70 pmol/L, respectively). Based on meta-analysis, the sensitivity is 74% and the specificity 80% for CA125. For HE4, the sensitivity is 73% and the specificity 79%.

CA125, HE4, CA15.3, and CA72.4 were the best to discriminate stage II-IV primary invasive from benign tumors, with AUCs between 0.85 (CA72.4) and 0.96 (HE4) (**Table 3**, **Supplementary** Figure 2). Of these four proteins, CA125 (0.78), HE4 (0.78), and CA72.4 (0.72) had AUCs above 0.7 to discriminate stage I primary invasive from benign tumors. AUCs to distinguish stage I invasive from borderline ovarian tumors were below 0.70 for all biomarkers (highest value 0.67). When subdividing these groups by histological subtype in a post hoc analysis, CA15.3, HE4, CA72.4 en Eotaxin-1 may have value for discriminating between borderline and serous epithelial ovarian stage I cancer (AUCs 0.73-0.85) (**Table 4**, **Supplementary** Figure 4).

### Adding proteins to clinical and ultrasound information to predict malignancy

The AUC of the seven ADNEX variables was 0.909 (95% CI 0.894-0.929) to discriminate between benign and malignant tumors (**Table 5**, **Supplementary** Figure 5). When proteins were individually added to this model, the highest improvement in AUC was observed for HE4 (AUC 0.935, 95% CI 0.922-0.949), CA125 (0.926, 95% CI 0.911-0.943), CA72.4 (0.923, 95%CI 0.908-0.943) and CA15.3 (0.916, 95% CI 0.901-0.933) (**Table 5**). Based on LASSO, seven proteins (HE4, CA72.4, CA125, IL6, Eotaxin2, Arginase, sFLT1) were selected on top of the ADNEX variables, resulting in an AUC of 0.935 (95% CI 0.927-0.955). The selection of these proteins was not consistent for the different bootstrap samples, except for HE4, CA125 and CA72.4 (**Supplementary** Figure 6). The prespecified combination of HE4 and CA125 resulted in an AUC of 0.937 (95% CI 0.925-0.951).

**Table 5.** Optimism-corrected AUC (95% Confidence interval) (benign vs malignant) and Polytomous Discrimination Index (95% Confidence interval).

| **Model** | **AUC**<br>**Benign vs Malignant** | **Polytomous**<br>**Discrimination Index** |
| --- | --- | --- |
| *ADNEX alone* | 0.909 (0.894-0.929) | 0.532 (0.510-0.589) |
| *ADNEX + 1 protein* |  |  |
| CA125 | 0.926 (0.911-0.943) | 0.581 (0.556-0.646) |
| HE4 | 0.935 (0.922-0.949) | 0.568 (0.547-0.636) |
| CA72.4 | 0.923 (0.908-0.943) | 0.551 (0.532-0.615) |
| CA15.3 | 0.916 (0.901-0.933) | 0.563 (0.541-0.621) |
| sFlt1 | 0.912 (0.896-0.930) | 0.530 (0.513-0.589) |
| MMP-7 | 0.911 (0.894-0.930) | 0.527 (0.514-0.590) |
| IL-6 | 0.911 (0.895-0.930) | 0.531 (0.511-0.595) |
| PIGF | 0.911 (0.896-0.930) | 0.522 (0.500-0.589) |
| proGRP | 0.910 (0.894-0.930) | 0.530 (0.510-0.588) |
| SAA | 0.910 (0.895-0.930) | 0.533 (0.511-0.595) |
| OPN | 0.910 (0.896-0.930) | 0.533 (0.514-0.592) |
| CA19.9 | 0.910 (0.894-0.929) | 0.533 (0.512-0.593) |
| MIG | 0.910 (0.894-0.930) | 0.531 (0.515-0.590) |
| CEA | 0.910 (0.895-0.929) | 0.546 (0.520-0.604) |
| Arginase | 0.910 (0.895-0.930) | 0.528 (0.508-0.586) |
| Eotaxin-2 | 0.910 (0.895-0.929) | 0.529 (0.507-0.594) |
| IP-10 | 0.910 (0.895-0.929) | 0.532 (0.511-0.595) |
| SDF-1alpha | 0.909 (0.894-0.929) | 0.526 (0.508-0.588) |
| VEGF-A | 0.909 (0.894-0.929) | 0.532 (0.512-0.595) |
| MMP-9 | 0.909 (0.894-0.929) | 0.527 (0.511-0.595) |
| LAP | 0.909 (0.894-0.929) | 0.526 (0.507-0.589) |
| IL-10 | 0.909 (0.894-0.930) | 0.535 (0.512-0.594) |
| Eotaxin-1 | 0.909 (0.894-0.929) | 0.528 (0.509-0.587) |
| RANTES | 0.909 (0.894-0.928) | 0.536 (0.514-0.600) |
| MDC | 0.909 (0.894-0.929) | 0.532 (0.508-0.592) |
| *ADNEX + proteins* |  |  |
| CA125 + HE4 | 0.937 (0.925-0.951) | 0.586 (0.559-0.649) |
| LASSO selection | 0.935 (0.927-0.955) | 0.565 (0.545-0.694) |

ADNEX, Assessment of Different NEoplasias in the adnexa model

### Adding proteins to clinical and ultrasound information to predict subtypes of malignancy

For the multiclass discrimination, the ADNEX variables had a PDI of 0.532 (95% CI 0.510- 0.589) (**Table 5**, **Supplementary** Figure 5). The highest improvement in PDI was observed when adding CA125 (0.581, 95% CI 0.556-0.646), HE4 (0.568, 95% CI 0.547-0.636), CA15.3 (0.563, 95% CI 0.541-0.621), CA72.4 (0.551, 95% CI 0.532-0.615) and CEA (0.546, 95% CI 0.520-0.604) (**Table 5**, **Supplementary** Figure 5). Selecting the best combination of proteins on top of the ADNEX variables retained four proteins (HE4, CA72.4, CA125 and CA15.3) with a PDI of 0.565 (95% CI 0.545-0.694). These proteins were selected with high certainty (**Supplementary** Figure 7). The prespecified combination of HE4 and CA125 resulted in a PDI of 0.586 (95% CI 0.559-0.649).

The analysis of pairwise AUCs indicated that the discrimination between stage II-IV primary invasive and secondary metastatic tumors benefited most from adding proteins to ADNEX (AUC increases up to 0.165), and the discrimination between benign and borderline tumors (increases at most 0.006) and between borderline and stage I primary invasive tumors (increases at most 0.016) least (**Table 6**). CA125 was particularly interesting to discriminate stage II-IV primary invasive from other tumors (AUC increases ranged between 0.019 and 0.165). HE4 was of most interest to discriminate benign from other tumors (increases between 0.006 and 0.028), and stage II-IV primary invasive from other malignant tumors (increases 0.026-0.132). CA72.4 had among the highest increases to discriminate secondary metastatic from other tumors except stage II-IV primary invasive (increases 0.029-0.034). CA15.3 had good results to discriminate borderline from primary invasive tumors (increases 0.014-0.032). MDC resulted in the highest increase 0.016) to discriminate borderline from stage I primary invasive tumors, and CEA in the highest increase (0.041) for stage I primary invasive vs secondary metastatic tumors.

**Table 6.** Increase in pairwise AUC (95% Confidence interval) compared with ADNEX.

| Protein | Benign vs<br>Borderline | Benign vs<br>Stage I | Benign vs<br>Stage II-IV | Benign vs<br>Metastatic | Borderline vs<br>Stage I | Borderline vs<br>Stage II-IV | Borderline vs<br>Metastatic | Stage I vs<br>Stage II-IV | Stage I vs<br>Metastatic | Stage II-IV vs<br>Metastatic |
| --- | --- | --- | --- | --- | --- | --- | --- | --- | --- | --- |
| *ADNEX alone* | 0.87 (0.84-0.90) | 0.91 (0.89-0.94) | 0.95 (0.94-0.97) | 0.91 (0.88-0.95) | 0.70 (0.66-0.78) | 0.92 (0.89-0.95) | 0.85 (0.82-0.91) | 0.83 (0.79-0.89) | 0.72 (0.67-0.84) | 0.69 (0.61-0.79) |
| *ADNEX + 1*<br>*protein* |  |  |  |  |  |  |  |  |  |  |
| CA125 | 0.88 (0.85-0.91) | 0.92 (0.90-0.95) | 0.98 (0.97-0.99) | 0.91 (0.88-0.95) | 0.70 (0.66-0.79) | 0.93 (0.91-0.96) | 0.86 (0.82-0.92) | 0.87 (0.83-0.92) | 0.74 (0.70-0.86) | 0.85 (0.82-0.91) |
| HE4 | 0.87 (0.85-0.91) | 0.94 (0.92-0.96) | 0.98 (0.97-0.99) | 0.93 (0.89-0.97) | 0.70 (0.65-0.79) | 0.94 (0.92-0.97) | 0.86 (0.82-0.92) | 0.86 (0.82-0.91) | 0.72 (0.68-0.85) | 0.82 (0.78-0.91) |
| CA72.4 | 0.87 (0.85-0.91) | 0.93 (0.91-0.95) | 0.97 (0.96-0.98) | 0.94 (0.91-0.97) | 0.71 (0.67-0.79) | 0.93 (0.90-0.96) | 0.89 (0.86-0.94) | 0.83 (0.79-0.89) | 0.75 (0.69-0.86) | 0.66 (0.62-0.78) |
| CA15.3 | 0.87 (0.84-0.90) | 0.92 (0.90-0.95) | 0.97 (0.96-0.98) | 0.91 (0.88-0.96) | 0.72 (0.68-0.80) | 0.95 (0.93-0.97) | 0.87 (0.83-0.92) | 0.84 (0.81-0.90) | 0.73 (0.68-0.84) | 0.79 (0.74-0.87) |
| sFlt1 | 0.87 (0.84-0.90) | 0.91 (0.89-0.94) | 0.96 (0.94-0.97) | 0.91 (0.88-0.95) | 0.71 (0.67-0.79) | 0.91 (0.89-0.94) | 0.85 (0.81-0.91) | 0.83 (0.80-0.89) | 0.72 (0.67-0.84) | 0.68 (0.63-0.78) |
| MMP-7 | 0.87 (0.85-0.90) | 0.91 (0.89-0.94) | 0.96 (0.94-0.97) | 0.92 (0.88-0.96) | 0.70 (0.66-0.77) | 0.92 (0.89-0.95) | 0.86 (0.82-0.92) | 0.83 (0.79-0.89) | 0.72 (0.69-0.84) | 0.67 (0.61-0.78) |
| IL-6 | 0.87 (0.84-0.90) | 0.91 (0.89-0.94) | 0.96 (0.94-0.97) | 0.91 (0.89-0.95) | 0.70 (0.66-0.78) | 0.92 (0.89-0.95) | 0.86 (0.82-0.92) | 0.83 (0.79-0.89) | 0.71 (0.66-0.84) | 0.69 (0.64-0.79) |
| PlGF | 0.87 (0.85-0.90) | 0.91 (0.89-0.94) | 0.96 (0.95-0.97) | 0.91 (0.87-0.96) | 0.70 (0.66-0.78) | 0.92 (0.90-0.95) | 0.85 (0.81-0.92) | 0.84 (0.80-0.89) | 0.72 (0.68-0.85) | 0.66 (0.61-0.79) |
| proGRP | 0.87 (0.84-0.90) | 0.91 (0.89-0.94) | 0.96 (0.94-0.97) | 0.91 (0.88-0.95) | 0.70 (0.65-0.78) | 0.91 (0.89-0.94) | 0.85 (0.81-0.91) | 0.83 (0.80-0.89) | 0.72 (0.66-0.84) | 0.67 (0.61-0.78) |
| SAA | 0.87 (0.84-0.90) | 0.91 (0.89-0.94) | 0.96 (0.94-0.97) | 0.91 (0.88-0.95) | 0.70 (0.66-0.78) | 0.92 (0.90-0.95) | 0.85 (0.82-0.91) | 0.84 (0.80-0.90) | 0.74 (0.68-0.86) | 0.68 (0.61-0.79) |
| OPN | 0.87 (0.84-0.90) | 0.91 (0.89-0.94) | 0.96 (0.94-0.97) | 0.91 (0.88-0.96) | 0.70 (0.65-0.78) | 0.92 (0.89-0.95) | 0.87 (0.84-0.92) | 0.83 (0.79-0.89) | 0.73 (0.69-0.85) | 0.70 (0.64-0.80) |
| CA19.9 | 0.87 (0.84-0.90) | 0.91 (0.89-0.94) | 0.95 (0.94-0.97) | 0.91 (0.88-0.95) | 0.71 (0.66-0.78) | 0.91 (0.89-0.94) | 0.85 (0.81-0.91) | 0.83 (0.80-0.89) | 0.74 (0.69-0.85) | 0.69 (0.63-0.79) |
| MIG | 0.87 (0.84-0.90) | 0.91 (0.89-0.94) | 0.96 (0.94-0.97) | 0.92 (0.89-0.95) | 0.70 (0.66-0.79) | 0.91 (0.89-0.94) | 0.87 (0.83-0.93) | 0.83 (0.80-0.89) | 0.74 (0.70-0.85) | 0.69 (0.63-0.79) |
| CEA | 0.87 (0.85-0.91) | 0.91 (0.89-0.94) | 0.95 (0.94-0.97) | 0.91 (0.88-0.96) | 0.69 (0.66-0.78) | 0.91 (0.89-0.94) | 0.86 (0.82-0.92) | 0.83 (0.79-0.89) | 0.76 (0.70-0.87) | 0.73 (0.67-0.82) |
| Arginase | 0.87 (0.85-0.90) | 0.91 (0.89-0.94) | 0.96 (0.94-0.97) | 0.91 (0.88-0.95) | 0.70 (0.65-0.78) | 0.91 (0.89-0.94) | 0.85 (0.82-0.91) | 0.83 (0.80-0.89) | 0.72 (0.67-0.84) | 0.68 (0.63-0.79) |
| Eotaxin-2 | 0.87 (0.85-0.90) | 0.91 (0.89-0.94) | 0.95 (0.94-0.97) | 0.91 (0.88-0.95) | 0.70 (0.66-0.78) | 0.91 (0.89-0.94) | 0.84 (0.81-0.91) | 0.83 (0.79-0.89) | 0.71 (0.66-0.84) | 0.67 (0.61-0.78) |
| IP-10 | 0.87 (0.84-0.90) | 0.91 (0.89-0.94) | 0.96 (0.95-0.97) | 0.91 (0.88-0.95) | 0.70 (0.66-0.78) | 0.93 (0.90-0.95) | 0.85 (0.81-0.91) | 0.84 (0.81-0.89) | 0.71 (0.67-0.84) | 0.68 (0.63-0.79) |
| SDF-1alpha | 0.87 (0.85-0.90) | 0.91 (0.89-0.94) | 0.95 (0.94-0.97) | 0.91 (0.88-0.95) | 0.70 (0.66-0.78) | 0.91 (0.89-0.94) | 0.86 (0.81-0.91) | 0.83 (0.79-0.89) | 0.71 (0.66-0.84) | 0.68 (0.62-0.79) |
| VEGF-A | 0.87 (0.85-0.90) | 0.91 (0.89-0.94) | 0.95 (0.94-0.97) | 0.91 (0.88-0.95) | 0.70 (0.66-0.78) | 0.92 (0.89-0.95) | 0.85 (0.81-0.92) | 0.83 (0.80-0.89) | 0.72 (0.68-0.85) | 0.68 (0.61-0.78) |
| MMP-9 | 0.87 (0.84-0.90) | 0.91 (0.90-0.94) | 0.95 (0.94-0.97) | 0.91 (0.88-0.95) | 0.70 (0.65-0.78) | 0.91 (0.89-0.94) | 0.85 (0.81-0.91) | 0.83 (0.79-0.89) | 0.73 (0.68-0.85) | 0.69 (0.63-0.79) |
| LAP | 0.87 (0.84-0.90) | 0.91 (0.89-0.94) | 0.95 (0.94-0.97) | 0.91 (0.88-0.95) | 0.70 (0.65-0.79) | 0.92 (0.90-0.95) | 0.86 (0.82-0.92) | 0.83 (0.79-0.89) | 0.72 (0.68-0.84) | 0.68 (0.62-0.78) |
| IL-10 | 0.87 (0.84-0.90) | 0.91 (0.89-0.94) | 0.95 (0.94-0.97) | 0.91 (0.88-0.95) | 0.71 (0.66-0.79) | 0.91 (0.89-0.94) | 0.85 (0.82-0.91) | 0.83 (0.80-0.89) | 0.73 (0.68-0.84) | 0.67 (0.62-0.79) |
| Eotaxin-1 | 0.87 (0.84-0.90) | 0.91 (0.89-0.94) | 0.96 (0.94-0.97) | 0.91 (0.88-0.95) | 0.71 (0.66-0.79) | 0.92 (0.89-0.95) | 0.86 (0.83-0.91) | 0.83 (0.79-0.89) | 0.71 (0.67-0.84) | 0.67 (0.60-0.78) |
| RANTES | 0.87 (0.84-0.90) | 0.91 (0.89-0.94) | 0.95 (0.94-0.97) | 0.91 (0.88-0.95) | 0.70 (0.66-0.78) | 0.91 (0.89-0.94) | 0.86 (0.83-0.92) | 0.83 (0.79-0.89) | 0.74 (0.69-0.85) | 0.71 (0.64-0.79) |
| MDC | 0.87 (0.84-0.90) | 0.91 (0.89-0.94) | 0.95 (0.94-0.97) | 0.91 (0.88-0.95) | 0.72 (0.67-0.80) | 0.91 (0.89-0.94) | 0.85 (0.81-0.91) | 0.83 (0.80-0.89) | 0.74 (0.68-0.85) | 0.68 (0.62-0.79) |
| *ADNEX + proteins* |  |  |  |  |  |  |  |  |  |  |
| CA125 + HE4 | 0.88 (0.85-0.91) | 0.94 (0.92-0.96) | 0.99 (0.98-0.99) | 0.92 (0.89-0.96) | 0.70 (0.66-0.79) | 0.95 (0.93-0.97) | 0.87 (0.83-0.92) | 0.87 (0.84-0.92) | 0.73 (0.70-0.86) | 0.86 (0.83-0.93) |
| LASSO | 0.87 (0.86-0.92) | 0.93 (0.92-0.96) | 0.98 (0.97-0.99) | 0.92 (0.91-0.97) | 0.69 (0.65-0.81) | 0.94 (0.92-0.97) | 0.86 (0.84-0.94) | 0.86 (0.85-0.92) | 0.73 (0.71-0.89) | 0.84 (0.82-0.93) |

ADNEX, Assessment of Different NEoplasias in the adnexa model

## Discussion

Our study supports the status of CA125 as the best serum protein to discriminate benign from malignant ovarian tumors, with an AUC of 0.84. However, when adding biomarkers to the ADNEX version without CA125 (i.e. only based on clinical and ultrasound parameters), HE4 increased the AUC more than CA125. For discrimination between five major tumor types, adding CA125 had the highest increase in PDI, followed by HE4. These two markers appeared particularly useful to identify stage II-IV primary invasive tumors. CA72.4 was promising as a biomarker to improve discrimination between secondary metastatic and non-invasive or early stage tumors, CA15.3 as a biomarker to improve discrimination between borderline and primary invasive tumors.

This is by far the largest study on (immune-related) biomarkers in combination with ultrasound for ovarian tumors. Our study also has some limitations. First, measuring proteins are snapshots and do not provide any functional information. Second, to avoid interference, we excluded patients with a prior history of cancer and immune-related diseases. While this refers to a small fraction of patients, they also need an appropriate diagnostic work-up in daily clinical practice. Third, most of patients have been recruited in centers with a high prevalence of malignancy. This increases the number of malignancies, which reduces uncertainty in the analysis for rarer subtypes (borderline, stage I primary invasive, secondary metastatic). Nevertheless, this tends to result in an overrepresentation of high risk patients. Despite this, the number of secondary metastatic tumors (n=31) remained modest. Given the number of proteins evaluated, and the low or modest amount of patients with borderline, stage I primary invasive, or secondary metastatic tumors, we labeled our study as exploratory.

Our results indicate that the preoperative discrimination of a borderline tumors versus a stage I ovarian cancer remains difficult, with univariable AUCs of at most 0.67 for all proteins. Nevertheless, subclassification of the masses would have a major clinical impact, since the prognosis is different in both patient groups. Moreover, patients with these tumors are often young women with a desire to retain their fertility, such that the choice between more extensive (stage I invasive) versus a more conservative (borderline tumor) surgical intervention is vital. However, when considering the specific histopathology of these tumors, some proteins emerged as potential biomarkers to differentiate serous borderline from serous epithelial stage I primary invasive tumors. These findings could be due to chance, but perhaps also to the different biological and immunological behavior of these tumors. The ADNEX variables alone resulted in an AUC of 0.705 to discriminate borderline from stage I primary invasive tumors. Addition of MDC or CA15.3 increased the AUC to 0.721 or 0.719, which is still suboptimal. Nevertheless, there is a biological rationale of MDC as a possible candidate biomarker for these two malignancy subtypes. MDC is being secreted by protumoral macrophages, leading to recruitment of regulatory T cells and T helper 2 cells, increasing the immune suppressive microenvironment.^27,28^ It can be expected that its expression is different between an invasive and a non-invasive neoplasm.

While the search for biomarkers to differentiate between types of ovarian tumors is important, we realize that the ADNEX variables (without CA125) already result in an AUC >0.9 to discriminate between benign and malignant tumors. This means that the room for improvement beyond these variables is limited. Nevertheless, this is partly an ethical and economical debate that goes beyond the scope of this study. All biomarkers come with a cost, are not accessible in all hospitals and certainly not in low-resource countries.

In conclusion, CA125 and HE4 are the most important biomarkers to diagnose ovarian tumors, either univariably or as addition to established clinical and ultrasound variables. Biomarkers that merit further study are CA72.4 and CA15.3, as these may be of use to improve the discrimination between malignant subtypes. The discrimination between borderline and stage I primary invasive tumors is not strongly improved by adding proteins, and remains a difficult yet important clinical problem.

## Data sharing statement

The pseudonymized data and data dictionary is stored in the KU Leuven Research Data Repository (RDR), https://doi.org/10.48804/TXL95Z. The dataset is not publicly available because this was not part of the informed consent. However, the dataset may be obtained following permission of prof. AC and prof. DT and after fulfilling all data transfer requirements.

