## Supplementary material for "Added value of serum proteins to clinical and ultrasound information in predicting the risk of malignancy in ovarian tumors"

#shared first; *shared last author

**Supplementary Appendix 1.** Isolation of serum.

Take one blood sample of 8.5 ml with a serum tube (BD Vacutainer SST II advance (ref 367953)) .

Ideally blood is centrifuged <u>the same day</u> but if not otherwise possible can be processed later on, by the latest after 2 days (if processed > 24h, please take note of it). Keep the blood tubes at room temperature.

- Centrifuge at 800 rcf, accel 9, decel 9, during 10 minutes at 20°C
- Collect supernatant (=serum) in non-sterile cryovials (VWR 720-1003)
  - Collection
    - First cryovials: 500 µl
    - Second cryovials: 200 µl
    - The rest (when possible) in 200 µl
    - The last: the remaining amount of serum.
  - Pseudonymized labelling of the sample
- Store at -80°C
- Frozen samples will be shipped once every three-five months on dry ice to UZ Leuven (Belgium)

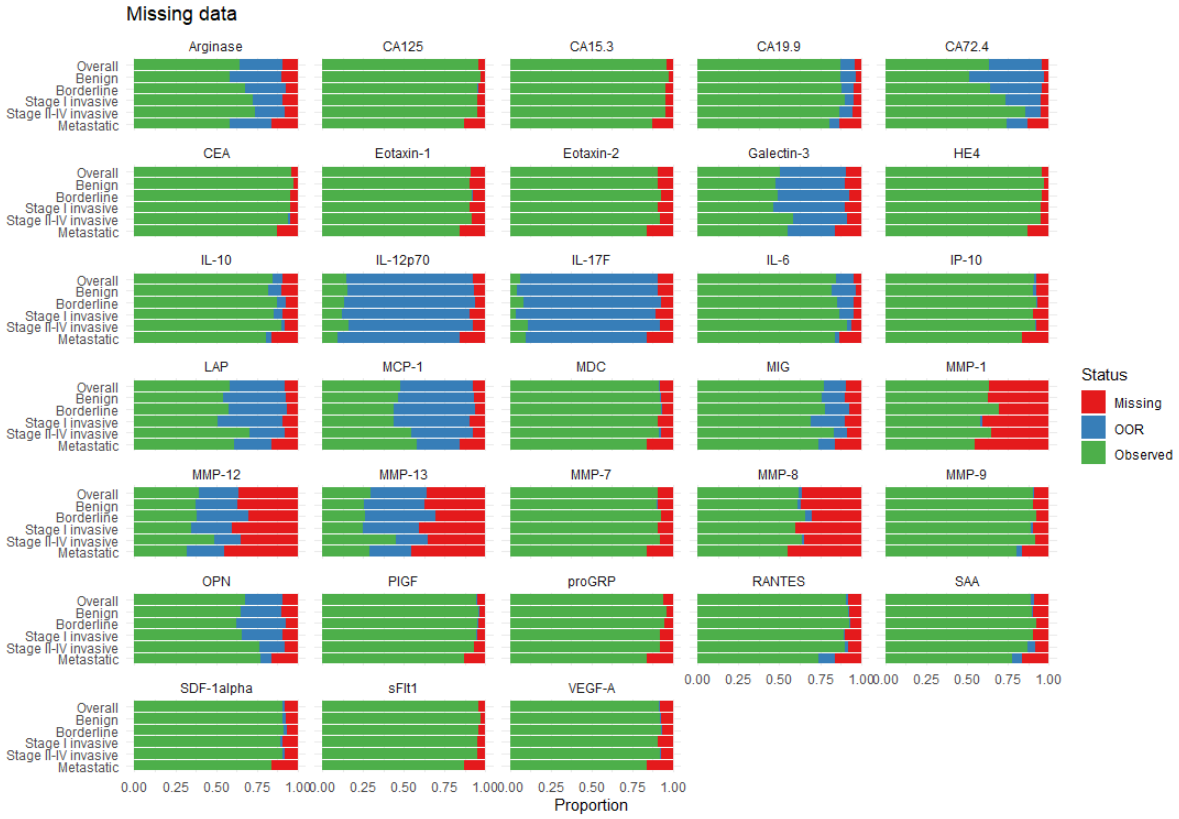


**Supplementary Figure 1.** Overview of proportion observed, out of range and missing observations.

**Supplementary Table 1.** Correlation between ADNEX-variables and the biomarkers.

| | Age | Maximum Diameter Lesion | Proportion solid tissue | > 10 locules | Number papillary projections | Acoustic shadows | Ascites |
|---|---|---|---|---|---|---|---|
| IL-10 | 0.09 | 0.01 | 0.13 | -0.02 | -0.07 | 0.05 | -0.08 |
| IP-10 | 0.18 | -0.07 | 0.16 | 0.05 | -0.05 | 0.11 | -0.19 |
| MIG | 0.28 | -0.01 | 0.23 | 0.03 | -0.06 | 0.04 | -0.01 |
| SDF-1alpha | -0.06 | -0.03 | -0.03 | 0.00 | 0.01 | 0.06 | -0.09 |
| VEGF-A | 0.13 | 0.01 | 0.12 | 0.05 | -0.12 | 0.16 | -0.27 |
| Arginase | 0.07 | 0 | 0.05 | -0.04 | -0.03 | 0.04 | -0.17 |
| LAP | 0.04 | 0.02 | 0.05 | 0.03 | -0.07 | 0.19 | -0.06 |
| MDC | -0.08 | -0.07 | 0.09 | 0.02 | -0.03 | -0.05 | -0.01 |
| RANTES | -0.08 | 0.01 | 0.04 | 0.02 | 0.02 | -0.04 | -0.03 |
| MMP-9 | -0.09 | -0.03 | 0.07 | 0.01 | -0.01 | 0.01 | -0.06 |
| Eotaxin-1 | 0.21 | -0.08 | 0.06 | 0.01 | -0.08 | 0.03 | 0.01 |
| Eotaxin-2 | 0.12 | -0.06 | 0.06 | 0.03 | -0.04 | 0.08 | 0.01 |
| MMP-7 | 0.34 | 0.11 | 0.22 | -0.02 | -0.03 | 0.15 | -0.26 |
| SAA | 0.16 | 0.11 | 0.21 | -0.01 | -0.03 | 0.10 | -0.33 |
| OPN | 0.13 | 0.13 | 0.18 | -0.03 | -0.04 | 0.02 | -0.33 |
| CEA | 0.32 | 0.10 | 0.06 | -0.15 | -0.01 | -0.01 | -0.11 |
| CA125 | 0.15 | 0.26 | 0.44 | 0.06 | -0.01 | 0.12 | -0.33 |
| PlGF | 0.54 | 0.16 | 0.28 | -0.03 | -0.07 | 0.09 | -0.32 |
| CA15.3 | 0.32 | 0.08 | 0.36 | -0.02 | -0.09 | 0.09 | -0.17 |
| sFlt1 | 0.34 | 0.09 | 0.18 | -0.02 | 0.03 | 0.06 | -0.13 |
| CA19.9 | 0.02 | 0.16 | -0.02 | -0.09 | 0 | -0.05 | -0.08 |
| IL-6 | 0.07 | 0.01 | 0.12 | 0.02 | -0.07 | 0.10 | -0.06 |
| CA72.4 | 0.24 | 0.12 | 0.30 | 0.01 | -0.10 | 0.11 | -0.18 |
| HE4 | 0.50 | 0.18 | 0.45 | 0.05 | -0.06 | 0.14 | -0.37 |
| proGRP | 0.37 | 0.09 | 0.07 | -0.02 | 0.02 | -0.01 | -0.13 |

The correlations with age, maximum diameter of lesion, proportion of solid tissue and number of papillary projections is calculated with the Spearman correlation. The correlations with presence of more than 10 locules, presence of acoustic shadows and presence of ascites is calculated with the point biserial correlation.

| | BENvMAL | BENvBOR | BENvST1 | BENvHST | BENvMET | BORvST1 | BORvHST | BORvMET | ST1vHST | ST1vMET | HSTvMET |
|---|---|---|---|---|---|---|---|---|---|---|---|
| CA125 | 0.84 | 0.74 | 0.78 | 0.95 | 0.72 | 0.58 | 0.85 | 0.52 | 0.80 | 0.56 | 0.83 |
| HE4 | 0.82 | 0.64 | 0.78 | 0.96 | 0.83 | 0.67 | 0.92 | 0.73 | 0.81 | 0.54 | 0.82 |
| CA72.4 | 0.75 | 0.60 | 0.72 | 0.85 | 0.83 | 0.62 | 0.77 | 0.77 | 0.67 | 0.68 | 0.52 |
| CA15.3 | 0.71 | 0.51 | 0.66 | 0.88 | 0.70 | 0.67 | 0.88 | 0.71 | 0.77 | 0.51 | 0.78 |
| PlGF | 0.66 | 0.53 | 0.60 | 0.76 | 0.70 | 0.57 | 0.73 | 0.66 | 0.67 | 0.60 | 0.57 |
| MMP-7 | 0.66 | 0.56 | 0.61 | 0.74 | 0.68 | 0.56 | 0.69 | 0.63 | 0.63 | 0.57 | 0.55 |
| IL-6 | 0.62 | 0.54 | 0.56 | 0.69 | 0.67 | 0.52 | 0.66 | 0.64 | 0.63 | 0.60 | 0.54 |
| SAA | 0.62 | 0.53 | 0.51 | 0.72 | 0.67 | 0.51 | 0.69 | 0.64 | 0.69 | 0.64 | 0.56 |
| sFlt1 | 0.61 | 0.57 | 0.57 | 0.66 | 0.62 | 0.51 | 0.59 | 0.55 | 0.58 | 0.54 | 0.54 |
| MIG | 0.61 | 0.52 | 0.51 | 0.69 | 0.70 | 0.50 | 0.70 | 0.71 | 0.68 | 0.70 | 0.51 |
| IP-10 | 0.60 | 0.51 | 0.52 | 0.71 | 0.58 | 0.53 | 0.72 | 0.59 | 0.68 | 0.56 | 0.63 |
| OPN | 0.59 | 0.52 | 0.58 | 0.65 | 0.70 | 0.59 | 0.66 | 0.70 | 0.57 | 0.61 | 0.54 |
| Arginase | 0.58 | 0.54 | 0.56 | 0.61 | 0.55 | 0.51 | 0.56 | 0.50 | 0.56 | 0.51 | 0.56 |
| VEGF-A | 0.58 | 0.52 | 0.51 | 0.66 | 0.66 | 0.53 | 0.68 | 0.67 | 0.65 | 0.64 | 0.52 |
| LAP | 0.57 | 0.51 | 0.50 | 0.63 | 0.65 | 0.51 | 0.63 | 0.65 | 0.63 | 0.64 | 0.51 |
| IL-10 | 0.57 | 0.52 | 0.51 | 0.63 | 0.62 | 0.51 | 0.61 | 0.61 | 0.62 | 0.63 | 0.51 |
| proGRP | 0.56 | 0.54 | 0.54 | 0.59 | 0.54 | 0.51 | 0.56 | 0.50 | 0.55 | 0.50 | 0.55 |
| CA19.9 | 0.55 | 0.53 | 0.63 | 0.53 | 0.53 | 0.59 | 0.50 | 0.51 | 0.59 | 0.61 | 0.51 |
| Eotaxin-1 | 0.55 | 0.55 | 0.54 | 0.61 | 0.57 | 0.60 | 0.66 | 0.62 | 0.56 | 0.52 | 0.55 |
| CEA | 0.53 | 0.53 | 0.56 | 0.51 | 0.67 | 0.53 | 0.54 | 0.64 | 0.57 | 0.61 | 0.67 |
| MMP-9 | 0.52 | 0.51 | 0.54 | 0.53 | 0.55 | 0.53 | 0.53 | 0.56 | 0.51 | 0.58 | 0.57 |
| RANTES | 0.52 | 0.52 | 0.51 | 0.53 | 0.59 | 0.52 | 0.51 | 0.61 | 0.53 | 0.59 | 0.62 |
| MDC | 0.52 | 0.52 | 0.58 | 0.55 | 0.58 | 0.60 | 0.53 | 0.55 | 0.64 | 0.66 | 0.52 |
| SDF-1alpha | 0.51 | 0.51 | 0.50 | 0.52 | 0.52 | 0.50 | 0.52 | 0.52 | 0.52 | 0.51 | 0.50 |
| Eotaxin-2 | 0.50 | 0.54 | 0.53 | 0.52 | 0.53 | 0.51 | 0.57 | 0.56 | 0.55 | 0.56 | 0.50 |

**Supplementary Figure 2.** Heatmap of the univariable AUC of each protein to discriminate between different tumor subgroups. Proteins are ordered according to the AUC for benign vs all malignant tumors (BEN vs MAL). BEN, benign tumor; MAL, all malignant tumors; BOR, borderline tumors; ST1, stage I primary invasive ovarian tumors; HST, stage II-IV primary invasive ovarian tumors; MET, secondary metastatic tumors.

**Supplementary Table 2.** AUC for the comparison of benign (n = 474) vs malignant (n = 458) per center and after meta-analysis of center-specific results (pooled AUC with 95% prediction interval).

| Protein | Leuven | Rome | Prague | London | Genk | Milan | AUC after MA (95% prediction interval) |
|---|---|---|---|---|---|---|---|
| CA125 | 0.84 (0.80; 0.88) | 0.83 (0.76; 0.88) | 0.82 (0.72; 0.88) | 0.88 (0.80; 0.93) | 0.78 (0.64; 0.88) | 0.83 (0.68; 0.91) | 0.83 (0.73; 0.90) |
| HE4 | 0.82 (0.77; 0.86) | 0.88 (0.81; 0.92) | 0.76 (0.66; 0.84) | 0.80 (0.71; 0.87) | 0.79 (0.65; 0.89) | 0.85 (0.72; 0.93) | 0.82 (0.67; 0.91) |
| CA72.4 | 0.75 (0.70; 0.80) | 0.74 (0.66;0.81) | 0.85 (0.76; 0.91) | 0.62 (0.51; 0.71) | 0.75 (0.60; 0.85) | 0.80 (0.65; 0.89) | 0.75 (0.47; 0.92) |
| CA15.3 | 0.68 (0.63; 0.73) | 0.76 (0.68; 0.82) | 0.70 (0.60; 0.79) | 0.64 (0.54; 0.74) | 0.73 (0.58; 0.84) | 0.79 (0.64; 0.89) | 0.71 (0.56; 0.83) |
| PlGF | 0.69 (0.64; 0.74) | 0.73 (0.65; 0.80) | 0.59 (0.48; 0.69) | 0.58 (0.47; 0.68) | 0.74 (0.59; 0.85) | 0.54 (0.38; 0.68) | 0.65 (0.38; 0.84) |
| MMP-7 | 0.71 (0.65; 0.76) | 0.68 (0.60; 0.75) | 0.62 (0.51; 0.72) | 0.57 (0.46; 0.67) | 0.55 (0.40; 0.69) | 0.62 (0.46; 0.75) | 0.64 (0.44; 0.80) |
| IL-6 | 0.67 (0.62; 0.72) | 0.61 (0.53; 0.69) | 0.64 (0.53; 0.74) | 0.56 (0.45; 0.66) | 0.73 (0.58; 0.84) | 0.67 (0.51; 0.80) | 0.64 (0.49; 0.78) |
| IP-10 | 0.61 (0.55; 0.66) | 0.65 (0.57; 0.73) | 0.62 (0.51; 0.72) | 0.57 (0.46; 0.67) | 0.66 (0.51; 0.78) | 0.62 (0.46; 0.75) | 0.62 (0.51; 0.72) |
| MIG | 0.62 (0.56; 0.67) | 0.65 (0.57; 0.73) | 0.62 (0.51; 0.72) | 0.54 (0.44; 0.65) | 0.56 (0.41; 0.70) | 0.69 (0.53; 0.81) | 0.61 (0.48; 0.73) |
| SAA | 0.61 (0.55; 0.67) | 0.64 (0.56; 0.72) | 0.55 (0.44; 0.66) | 0.58 (0.47; 0.68) | 0.58 (0.43; 0.72) | 0.70 (0.54; 0.82) | 0.61 (0.49; 0.73) |
| OPN | 0.60 (0.54; 0.65) | 0.67 (0.59; 0.75) | 0.64 (0.53; 0.73) | 0.56 (0.45; 0.66) | 0.54 (0.39; 0.68) | 0.71 (0.55; 0.83) | 0.62 (0.46; 0.76) |
| IL-10 | 0.56 (0.50; 0.62) | 0.66 (0.58; 0.74) | 0.57 (0.46; 0.68) | 0.63 (0.52; 0.72) | 0.51 (0.36; 0.65) | 0.71 (0.55; 0.83) | 0.60 (0.42; 0.76) |
| sFlt1 | 0.65 (0.59; 0.70) | 0.59 (0.51; 0.67) | 0.53 (0.42; 0.64) | 0.51 (0.40; 0.61) | 0.66 (0.51; 0.78) | 0.64 (0.48; 0.77) | 0.60 (0.41; 0.76) |
| Arginase | 0.58 (0.52; 0.64) | 0.63 (0.54; 0.71) | 0.56 (0.45; 0.66) | 0.54 (0.44; 0.65) | 0.56 (0.41; 0.70) | 0.70 (0.54; 0.82) | 0.59 (0.46; 0.72) |
| VEGF-A | 0.57 (0.52; 0.63) | 0.61 (0.53; 0.69) | 0.56 (0.45; 0.67) | 0.59 (0.49; 0.69) | 0.55 (0.41; 0.69) | 0.53 (0.37; 0.68) | 0.58 (0.47; 0.67) |
| LAP | 0.57 (0.51; 0.62) | 0.62 (0.54; 0.70) | 0.58 (0.47; 0.69) | 0.53 (0.42; 0.63) | 0.53 (0.38; 0.67) | 0.53 (0.38; 0.68) | 0.57 (0.45; 0.67) |
| proGRP | 0.60 (0.54; 0.66) | 0.58 (0.49; 0.66) | 0.53 (0.42; 0.64) | 0.52 (0.41; 0.62) | 0.54 (0.39; 0.68) | 0.59 (0.43; 0.73) | 0.57 (0.45; 0.68) |
| MDC | 0.50 (0.44; 0.56) | 0.61 (0.52; 0.69) | 0.57 (0.46; 0.67) | 0.53 (0.42; 0.63) | 0.67 (0.52; 0.79) | 0.56 (0.40; 0.70) | 0.56 (0.40; 0.73) |
| Eotaxin-1 | 0.57 (0.51; 0.63) | 0.53 (0.44; 0.61) | 0.60 (0.49; 0.70) | 0.57 (0.46; 0.67) | 0.50 (0.36; 0.65) | 0.64 (0.48; 0.77) | 0.57 (0.46; 0.68) |
| CEA | 0.55 (0.49; 0.61) | 0.51 (0.43; 0.60) | 0.60 (0.49; 0.70) | 0.57 (0.47; 0.67) | 0.71 (0.56; 0.82) | 0.51 (0.36; 0.66) | 0.57 (0.40; 0.72) |
| CA19.9 | 0.58 (0.52; 0.63) | 0.53 (0.44; 0.61) | 0.56 (0.45; 0.67) | 0.56 (0.45; 0.66) | 0.64 (0.49; 0.76) | 0.52 (0.36; 0.67) | 0.56 (0.45; 0.67) |
| MMP-9 | 0.55 (0.49; 0.60) | 0.56 (0.47; 0.64) | 0.50 (0.39; 0.61) | 0.52 (0.41; 0.62) | 0.60 (0.45; 0.73) | 0.58 (0.43; 0.73) | 0.55 (0.44; 0.65) |
| SDF-1α | 0.51 (0.45; 0.57) | 0.52 (0.43; 0.60) | 0.57 (0.46; 0.67) | 0.51 (0.41; 0.62) | 0.53 (0.38; 0.67) | 0.64 (0.48; 0.77) | 0.53 (0.42; 0.65) |
| Eotaxin-2 | 0.51 (0.45; 0.57) | 0.50 (0.42; 0.59) | 0.54 (0.43; 0.65) | 0.58 (0.47; 0.68) | 0.60 (0.45; 0.73) | 0.58 (0.43; 0.73) | 0.53 (0.42; 0.66) |
| RANTES | 0.53 (0.48; 0.59) | 0.51 (0.42; 0.59) | 0.51 (0.40; 0.61) | 0.53 (0.42; 0.64) | 0.51 (0.37; 0.66) | 0.50 (0.35; 0.65) | 0.52 (0.42; 0.61) |

MA, meta-analysis.

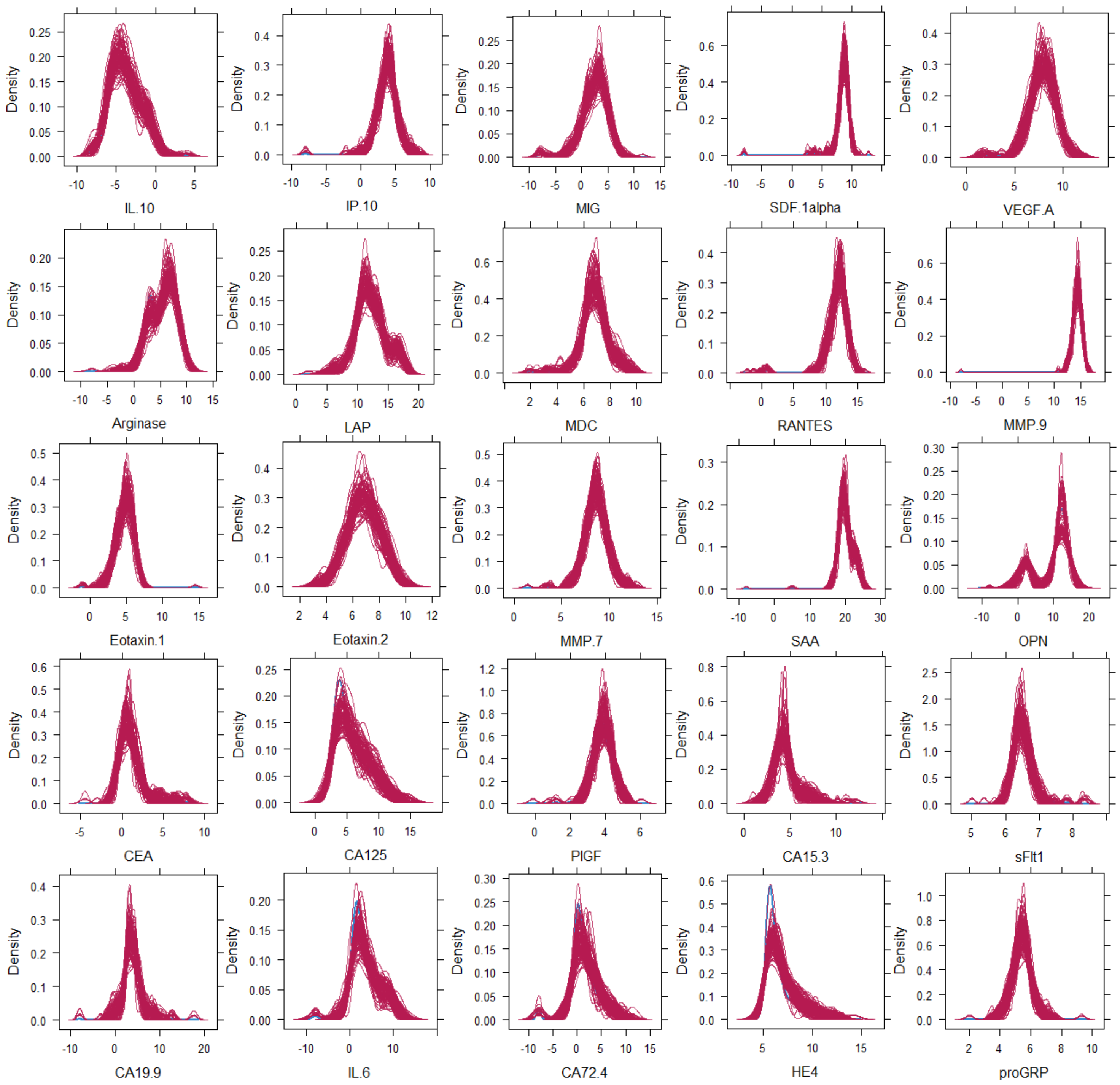


**Supplementary Figure 3.** Density plot of the proteins. The blue curve is the density of the values that were observed, the red curves are the densities for each of the 100 imputations of the values that were missing. The protein measurements on the x-axis are shown on logarithmic scale ($log_2$ transformation).

**Supplementary Table 3.** AUC for the comparison of benign (n = 474) vs malignant (n = 458) after multiple imputation (m = 100).

| Protein | Benign vs Malignant |
|---|---|
| CA125 | 0.84 (0.81; 0.87) |
| HE4 | 0.82 (0.79; 0.85) |
| CA72.4 | 0.75 (0.72; 0.78) |
| CA15.3 | 0.72 (0.68; 0.75) |
| PlGF | 0.66 (0.62; 0.69) |
| MMP-7 | 0.66 (0.62; 0.69) |
| IL-6 | 0.62 (0.59; 0.66) |
| IP-10 | 0.59 (0.55; 0.63) |
| MIG | 0.60 (0.56; 0.64) |
| SAA | 0.63 (0.59; 0.66) |
| OPN | 0.59 (0.55; 0.62) |
| IL-10 | 0.58 (0.55; 0.62) |
| sFlt1 | 0.61 (0.58; 0.65) |
| Arginase | 0.59 (0.55; 0.62) |
| VEGF-A | 0.57 (0.54; 0.61) |
| LAP | 0.57 (0.53; 0.60) |
| proGRP | 0.55 (0.52; 0.59) |
| MDC | 0.52 (0.48; 0.56) |
| Eotaxin-1 | 0.53 (0.49; 0.57) |
| CEA | 0.53 (0.49; 0.56) |
| CA19.9 | 0.55 (0.51; 0.59) |
| MMP-9 | 0.53 (0.49; 0.57) |
| SDF-1α | 0.51 (0.47; 0.54) |
| Eotaxin-2 | 0.51 (0.47; 0.55) |
| RANTES | 0.52 (0.48; 0.55) |

**Supplementary Table 4.** Sensitivity and specificity for CA125 and HE4 at common cut-offs. The results for both the meta-analysis and pooled analysis are shown.

| Center | CA125 (cut-off: 35 U/mL) | | HE4 (cut-off: 70 pmol/L) | |
|---|---|---|---|---|
| | Sensitivity (95% CI) | Specificity (95% CI) | Sensitivity (95% CI) | Specificity (95% CI) |
| Leuven | 70.3% (63.1-76.6) | 84.3% (78.6-88.7) | 73.1% (66.1-79.2) | 79.8% (73.7-84.8) |
| Prague | 73.4% (63.7-81.3) | 78.4% (62.8-88.6) | 72.3% (62.6-80.4) | 67.6% (51.5-80.4) |
| London | 78.4% (65.4-87.5) | 80.6% (69.1-88.6) | 66.7% (53.0-78.0) | 80.6% (69.1-88.6) |
| Genk | 47.4% (27.3-68.3) | 85.0% (73.9-91.9) | 63.2% (41.0-80.9) | 78.3% (66.4-86.9) |
| Milan | 82.5% (68.1-91.3) | 70.0% (48.1-85.5) | 77.5% (62.5-87.7) | 75.0% (53.1-88.8) |
| Rome | 78.5% (68.2-86.1) | 74.2% (64.7-81.9) | 78.5% (68.2-86.1) | 84.5% (76.0-90.4) |
| Meta-analysis | 73.8% (67.0-79.7) | 80.0% (74.3-84.7) | 73.0% (68.7-76.9) | 79.3% (75.3-82.7) |
| Pooled analysis | 73.4% (69.1-77.2) | 80.8% (77.0-84.1) | 73.1% (68.9-77.0) | 79.5% (75.7-82.9) |

| | BORvST1 | BORvsHGSOC | sBORvHGSOC | nsBORvnsEPI | BORvNonEPI |
|---|---|---|---|---|---|
| CA125 | 0.58 | 0.66 | 0.62 | 0.62 | 0.50 |
| HE4 | 0.67 | 0.81 | 0.82 | 0.66 | 0.51 |
| CA72.4 | 0.62 | 0.73 | 0.77 | 0.61 | 0.59 |
| CA15.3 | 0.67 | 0.85 | 0.89 | 0.63 | 0.56 |
| PlGF | 0.57 | 0.53 | 0.54 | 0.56 | 0.52 |
| MMP-7 | 0.56 | 0.55 | 0.58 | 0.57 | 0.63 |
| IL-6 | 0.52 | 0.51 | 0.50 | 0.55 | 0.53 |
| SAA | 0.51 | 0.52 | 0.52 | 0.50 | 0.57 |
| sFlt1 | 0.51 | 0.51 | 0.50 | 0.50 | 0.51 |
| MIG | 0.50 | 0.66 | 0.65 | 0.52 | 0.52 |
| IP-10 | 0.53 | 0.69 | 0.68 | 0.52 | 0.56 |
| OPN | 0.59 | 0.57 | 0.59 | 0.57 | 0.62 |
| Arginase | 0.51 | 0.62 | 0.63 | 0.53 | 0.53 |
| VEGF-A | 0.53 | 0.56 | 0.57 | 0.53 | 0.53 |
| LAP | 0.51 | 0.66 | 0.63 | 0.50 | 0.52 |
| IL-10 | 0.51 | 0.53 | 0.55 | 0.55 | 0.50 |
| proGRP | 0.51 | 0.54 | 0.51 | 0.50 | 0.56 |
| CA19.9 | 0.59 | 0.57 | 0.51 | 0.57 | 0.52 |
| Eotaxin-1 | 0.60 | 0.73 | 0.73 | 0.61 | 0.55 |
| CEA | 0.53 | 0.59 | 0.52 | 0.51 | 0.58 |
| MMP-9 | 0.53 | 0.55 | 0.58 | 0.60 | 0.50 |
| RANTES | 0.52 | 0.64 | 0.65 | 0.51 | 0.53 |
| MDC | 0.60 | 0.63 | 0.66 | 0.56 | 0.64 |
| SDF-1alpha | 0.50 | 0.61 | 0.59 | 0.51 | 0.53 |
| Eotaxin-2 | 0.51 | 0.67 | 0.66 | 0.53 | 0.66 |

**Supplementary Figure 4.** Heatmap representation of the univariable AUC of each protein to discriminate between different borderline and stage I invasive ovarian cancer. Results for comparison of all borderline tumors (BOT) with all stage I invasive tumors (ST1), the comparison of sereus BOT (sBOT) with high grade serous stage I (HGSOC), the comparison of non-serous BOT (nsBOT) with non-serous epithelial invasive ovarian cancer stage I (nsEPI) and the comparison of all BOT with non-epithelial ovarian cancers stage I (nonEPI).

| | BENvMAL | BENvBOR | BENvST1 | BENvHST | BENvMET | BORvST1 | BORvHST | BORvMET | ST1vHST | ST1vMET | HSTvMET | PDI |
|---|---|---|---|---|---|---|---|---|---|---|---|---|
| CA125 | 0.926 | 0.875 | 0.921 | 0.976 | 0.914 | 0.698 | 0.935 | 0.855 | 0.868 | 0.741 | 0.851 | 0.581 |
| :A125 + HE4 | 0.937 | 0.877 | 0.936 | 0.986 | 0.925 | 0.704 | 0.947 | 0.865 | 0.872 | 0.731 | 0.862 | 0.586 |
| LASSO | 0.935 | 0.870 | 0.927 | 0.980 | 0.923 | 0.691 | 0.943 | 0.862 | 0.855 | 0.729 | 0.841 | 0.565 |
| HE4 | 0.935 | 0.875 | 0.937 | 0.982 | 0.926 | 0.701 | 0.943 | 0.861 | 0.858 | 0.721 | 0.818 | 0.568 |
| CA72.4 | 0.923 | 0.870 | 0.929 | 0.969 | 0.939 | 0.711 | 0.926 | 0.888 | 0.833 | 0.755 | 0.664 | 0.551 |
| CA15.3 | 0.916 | 0.868 | 0.918 | 0.969 | 0.912 | 0.719 | 0.948 | 0.867 | 0.845 | 0.729 | 0.787 | 0.563 |
| sFlt1 | 0.912 | 0.868 | 0.911 | 0.956 | 0.913 | 0.710 | 0.914 | 0.852 | 0.831 | 0.720 | 0.678 | 0.530 |
| MMP-7 | 0.911 | 0.867 | 0.912 | 0.957 | 0.916 | 0.699 | 0.918 | 0.860 | 0.832 | 0.717 | 0.671 | 0.527 |
| IL-6 | 0.911 | 0.868 | 0.913 | 0.955 | 0.914 | 0.700 | 0.918 | 0.857 | 0.831 | 0.710 | 0.685 | 0.531 |
| PIGF | 0.911 | 0.868 | 0.911 | 0.959 | 0.910 | 0.702 | 0.922 | 0.850 | 0.836 | 0.715 | 0.664 | 0.522 |
| proGRP | 0.910 | 0.868 | 0.910 | 0.955 | 0.909 | 0.701 | 0.914 | 0.851 | 0.830 | 0.717 | 0.667 | 0.530 |
| SAA | 0.910 | 0.868 | 0.911 | 0.957 | 0.911 | 0.700 | 0.924 | 0.851 | 0.836 | 0.737 | 0.679 | 0.533 |
| OPN | 0.910 | 0.868 | 0.911 | 0.956 | 0.914 | 0.700 | 0.917 | 0.868 | 0.826 | 0.729 | 0.696 | 0.533 |
| CA19.9 | 0.910 | 0.867 | 0.914 | 0.954 | 0.910 | 0.705 | 0.914 | 0.853 | 0.830 | 0.736 | 0.688 | 0.533 |
| MIG | 0.910 | 0.869 | 0.909 | 0.956 | 0.918 | 0.700 | 0.915 | 0.874 | 0.830 | 0.738 | 0.686 | 0.531 |
| CEA | 0.910 | 0.873 | 0.911 | 0.953 | 0.911 | 0.694 | 0.913 | 0.865 | 0.827 | 0.762 | 0.730 | 0.546 |
| Arginase | 0.910 | 0.868 | 0.911 | 0.956 | 0.909 | 0.699 | 0.913 | 0.852 | 0.830 | 0.719 | 0.678 | 0.528 |
| Eotaxin-2 | 0.910 | 0.869 | 0.910 | 0.954 | 0.908 | 0.700 | 0.914 | 0.843 | 0.830 | 0.709 | 0.670 | 0.529 |
| IP-10 | 0.910 | 0.868 | 0.911 | 0.958 | 0.909 | 0.705 | 0.926 | 0.854 | 0.839 | 0.712 | 0.682 | 0.532 |
| SDF-1alpha | 0.909 | 0.870 | 0.911 | 0.954 | 0.909 | 0.704 | 0.915 | 0.856 | 0.827 | 0.715 | 0.683 | 0.526 |
| VEGF-A | 0.909 | 0.869 | 0.911 | 0.954 | 0.908 | 0.702 | 0.916 | 0.853 | 0.832 | 0.725 | 0.676 | 0.532 |
| MMP-9 | 0.909 | 0.868 | 0.913 | 0.954 | 0.911 | 0.704 | 0.915 | 0.851 | 0.830 | 0.735 | 0.694 | 0.527 |
| LAP | 0.909 | 0.868 | 0.911 | 0.954 | 0.910 | 0.705 | 0.921 | 0.863 | 0.829 | 0.719 | 0.677 | 0.526 |
| IL-10 | 0.909 | 0.866 | 0.911 | 0.953 | 0.909 | 0.709 | 0.915 | 0.854 | 0.831 | 0.726 | 0.670 | 0.535 |
| Eotaxin-1 | 0.909 | 0.868 | 0.910 | 0.955 | 0.911 | 0.707 | 0.919 | 0.858 | 0.830 | 0.711 | 0.673 | 0.528 |
| RANTES | 0.909 | 0.868 | 0.910 | 0.953 | 0.909 | 0.700 | 0.915 | 0.862 | 0.829 | 0.735 | 0.709 | 0.536 |
| MDC | 0.909 | 0.866 | 0.913 | 0.954 | 0.908 | 0.721 | 0.913 | 0.852 | 0.834 | 0.739 | 0.682 | 0.532 |
| ADNEX | 0.909 | 0.869 | 0.911 | 0.954 | 0.910 | 0.705 | 0.916 | 0.855 | 0.832 | 0.721 | 0.686 | 0.532 |

**Supplementary Figure 5.** Heatmap of the AUC and PDI for the added diagnostic value of the proteins. ADNEX (at the bottom) is the baseline model with only clinical and ultrasound variables. The other lines show the performance of the baseline model in combination with the biomarker(s). Proteins selected with LASSO are HE4, CA72.4, CA125, IL6, Eotaxin2, Arginase and sFLT1 for the discrimination between benign and malignant. HE4, CA72.4, CA125 and CA15.3 were selected for the other comparisons. BEN, benign tumor; MAL, all malignant tumors; BOR, borderline tumors; ST1, stage I primary invasive ovarian tumors; HST, stage II-IV primary invasive ovarian tumors; MET, secondary metastatic tumors; PDI, polytomous discrimination index.

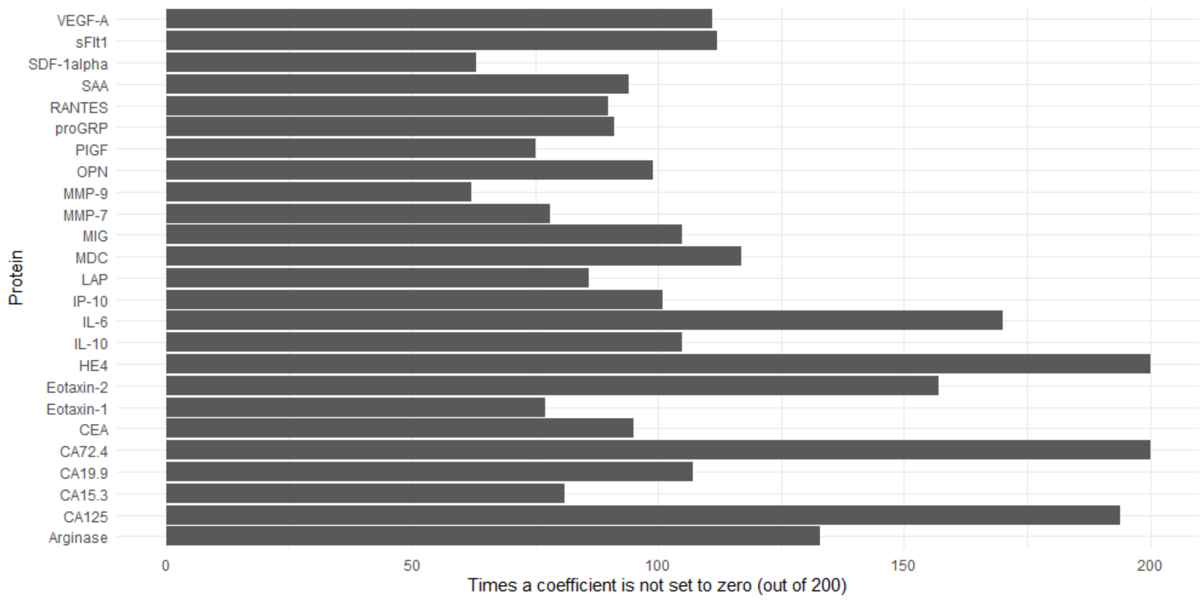


**Supplementary Figure 6.** Visualisation of the selection frequency across the bootstrap samples for the logistic model of benign (n = 474) vs malignant (n = 458).

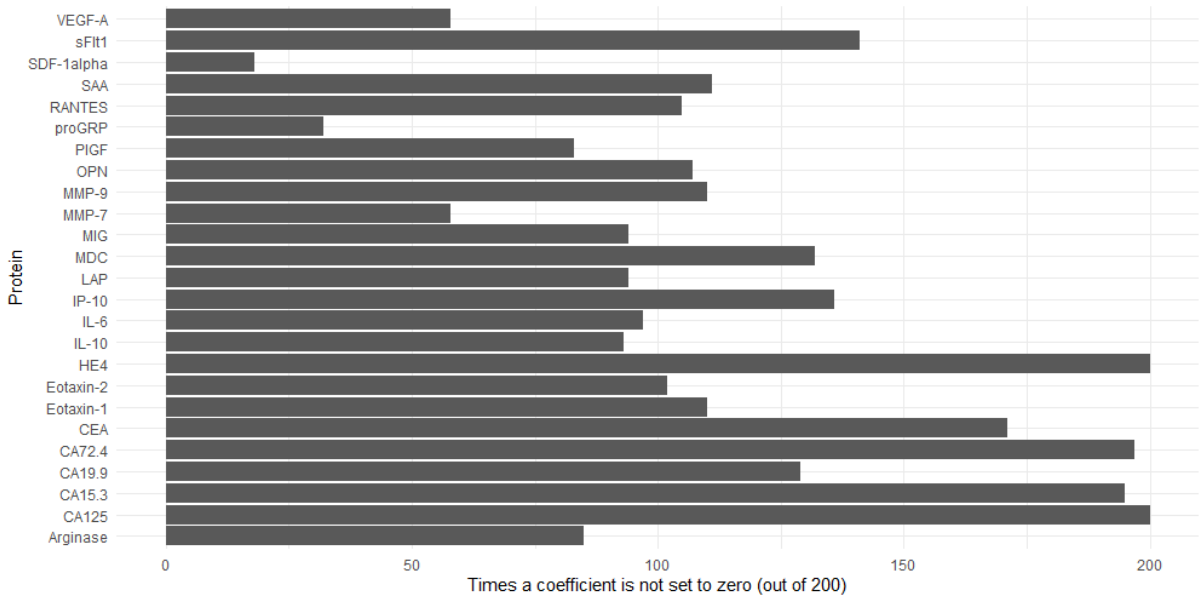


**Supplementary Figure 7.** Visualisation of the selection frequency across the bootstrap samples for the multinomial model.
